# Association of Common and Rare Variants with Alzheimer’s Disease in over 13,000 Diverse Individuals with Whole-Genome Sequencing from the Alzheimer’s Disease Sequencing Project

**DOI:** 10.1101/2023.09.01.23294953

**Authors:** Wan-Ping Lee, Seung Hoan Choi, Margaret G Shea, Po-Liang Cheng, Beth A Dombroski, Achilleas N Pitsillides, Nancy L Heard-Costa, Hui Wang, Katia Bulekova, Amanda B Kuzma, Yuk Yee Leung, John J Farrell, Honghuang Lin, Adam Naj, Elizabeth E Blue, Frederick Nusetor, Dongyu Wang, Eric Boerwinkle, William S Bush, Xiaoling Zhang, Philip L De Jager, Josée Dupuis, Lindsay A Farrer, Myriam Fornage, Eden Martin, Margaret Pericak-Vance, Sudha Seshadri, Ellen M Wijsman, Li-San Wang, Gerard D Schellenberg, Anita L Destefano, Jonathan L Haines, Gina M Peloso

## Abstract

Alzheimer’s Disease (AD) is a common disorder of the elderly that is both highly heritable and genetically heterogeneous. Here, we investigated the association between AD and both common variants and aggregates of rare coding and noncoding variants in 13,371 individuals of diverse ancestry with whole genome sequence (WGS) data. Pooled-population analyses identified genetic variants in or near *APOE, BIN1*, and *LINC00320* significantly associated with AD (p < 5×10^-8^). Population-specific analyses identified a haplotype on chromosome 14 including *PSEN1* associated with AD in Hispanics, further supported by aggregate testing of rare coding and noncoding variants in this region. Finally, we observed suggestive associations (p < 5×10^-5^) of aggregates of rare coding rare variants in *ABCA7* among non-Hispanic Whites (p=5.4×10^-6^), and rare noncoding variants in the promoter of *TOMM40* distinct of *APOE* in pooled-population analyses (p=7.2×10^-8^). Complementary pooled-population and population-specific analyses offered unique insights into the genetic architecture of AD.

## Introduction

Alzheimer’s Disease (AD) is a progressive neurological disorder characterized by a decline in cognitive and memory functions, which ultimately results in the inability to carry out daily activities. It is the most common cause of dementia, affecting more than 50 million people worldwide. This number is projected to almost triple by 2050, reaching 152 million people, as the baby-boom generation (born between 1946 and 1964) has already begun to reach age 65 years and beyond^1^. The disease is more common among older individuals, with the risk increasing significantly after the age of 65 years. In the United States, it is estimated that around 6.5 million people currently have AD, with a projection of nearly 14 million by 2050^1^.

Although the underlying multidimensional causes of AD are not fully understood, evidence suggests that genetics plays a crucial role in the development of the disease. Rare coding changes in *PSEN1*^2^, *PSEN2*^3^, and *APP*^4–7^ underlie autosomal dominant early-onset AD, while other coding changes in these genes are associated with increased risk of late-onset AD. The *APOE* gene is the strongest susceptibility gene associated with AD^8,9^, with isoforms defined by common missense variants associated with large effects on AD risk. Individuals with one copy of the *APOE* ε4 allele have approximately a three-fold increased risk of developing AD, while those with two copies of the ε4 allele are at approximately a 12-fold increased risk^10^. The presence of the *APOE* ε4 allele is also associated with an earlier onset of AD^11^. Each of these loci were first identified by family-based studies.

Recent large-scale genome-wide association studies using array-based genotyping and imputation have identified more than 80 common genetic loci associated with AD^12–14^. It is estimated that 25% of phenotypic variation in AD remains unexplained by known genetic variants associated with AD^15,16^, suggesting that additional risk loci are yet to be discovered. While genotype arrays are a useful tool for studying genetic variants associated with AD, they have limitations when it comes to discovering rare or novel genetic variant associations with disease. Array-based genotyping relies on a pre-designed set of probes that target specific genetic loci across the genome. Computational imputation may mitigate this limitation and improve the accuracy and coverage of array-based genotype data. However, imputation accuracy is directly determined by the size and quality of the reference panel and observed array data used, as well as the underlying patterns of genetic variation in the populations being studied. In contrast, whole genome sequencing (WGS) enables a full-spectrum exploration of short insertion/deletions (INDELs) and single nucleotide variants (SNVs) across the genome and provides a comprehensive view of an individual’s genetic information, allowing for the testing of both common and rare genetic variants that may be unique to individuals or populations not previously observed.

The Alzheimer’s Disease Sequencing Project (ADSP) is a collaborative research effort that utilizes WGS to identify both protective and risk genetic factors for AD. Data are collected from diverse individuals, recognizing that genetic risk, incidence, and prevalence rates can vary across populations. For example, *APOE* ε4 is more common in White individuals (∼24%) and African Americans (∼26%) compared to Asian individuals (∼12%) or Hispanic individuals (∼15%) and the effect of *APOE* ε4 varies among populations^17,18^. The odds ratios of ε3/ε4 are 2.49 and 3.83 for African ancestry and European ancestry while ε4/ε4 are 8.17 and 14.35^19^. Leveraging large-scale WGS from the ADSP, we performed association testing of single variants (minor allele frequency [MAF] > 0.5%) as well as aggregates of rare (MAF < 1%) coding and non-coding variants in up to 13,371 individuals (N_cases_=6,519 and N_control_=6,852).

## Results

### Overview

We performed association analyses using 13,371 of the 16,905 individuals in the ADSP Release 3 data to discover common and rare genetic variants associated with AD. The individuals that were excluded from analysis represent technical replicates, unexpected duplicates, and individuals with unknown AD status. Samples were sequenced by multiple centers with different platforms and libraries (**Table S1**). The Genome Center for Alzheimer’s Disease (GCAD) mapped short reads against the reference genome hg38 using BWA MEM^20^, called variants using the GATK^21^ HaplotypeCaller for each sample, and then performed joint genotyping across all samples using GATK^22^. The GCAD quality control (QC) working group performed quality checks of variants and genotypes and assigned a quality annotation^23^.

A total of 13,371 individuals were available for association analysis with AD status available. In the 13,371 individuals, 177.6 million bi-allelic SNVs and 12.9 million bi-allelic INDELs were observed. Given the ADSP data is composed of diverse individuals, we performed association testing across all participants (pooled samples, N_cases_=6,519 and N_control_=6,852) and within the three subgroups: African Americans (AA, N_cases_=1,137 and N_control_=1,707), Hispanics (HIS, N_cases_=1,021 and N_control_=1,988), and Non-Hispanic White (NHW, N_cases_=4,230 and N_control_=3,109) defined by reported race and ethnicity (**Figure 1**; **Table 1**). We performed single variant association testing (MAF > 0.5%) as well as association testing of aggregates of rare (MAF < 1%) coding and non-coding variants within the pooled samples and each of the three subgroups (AA, HIS, and NHW). The pooled samples analysis is most powerful to detect associations when there are similar effects across subgroups while the subgroup-specific analyses are beneficial to detect subgroup-specific effects. We limited the analyses to bi-allelic SNVs and INDELs after preliminary analyses showed false-positive associations across the genome for multi-allelic variants.

**Figure 1.**
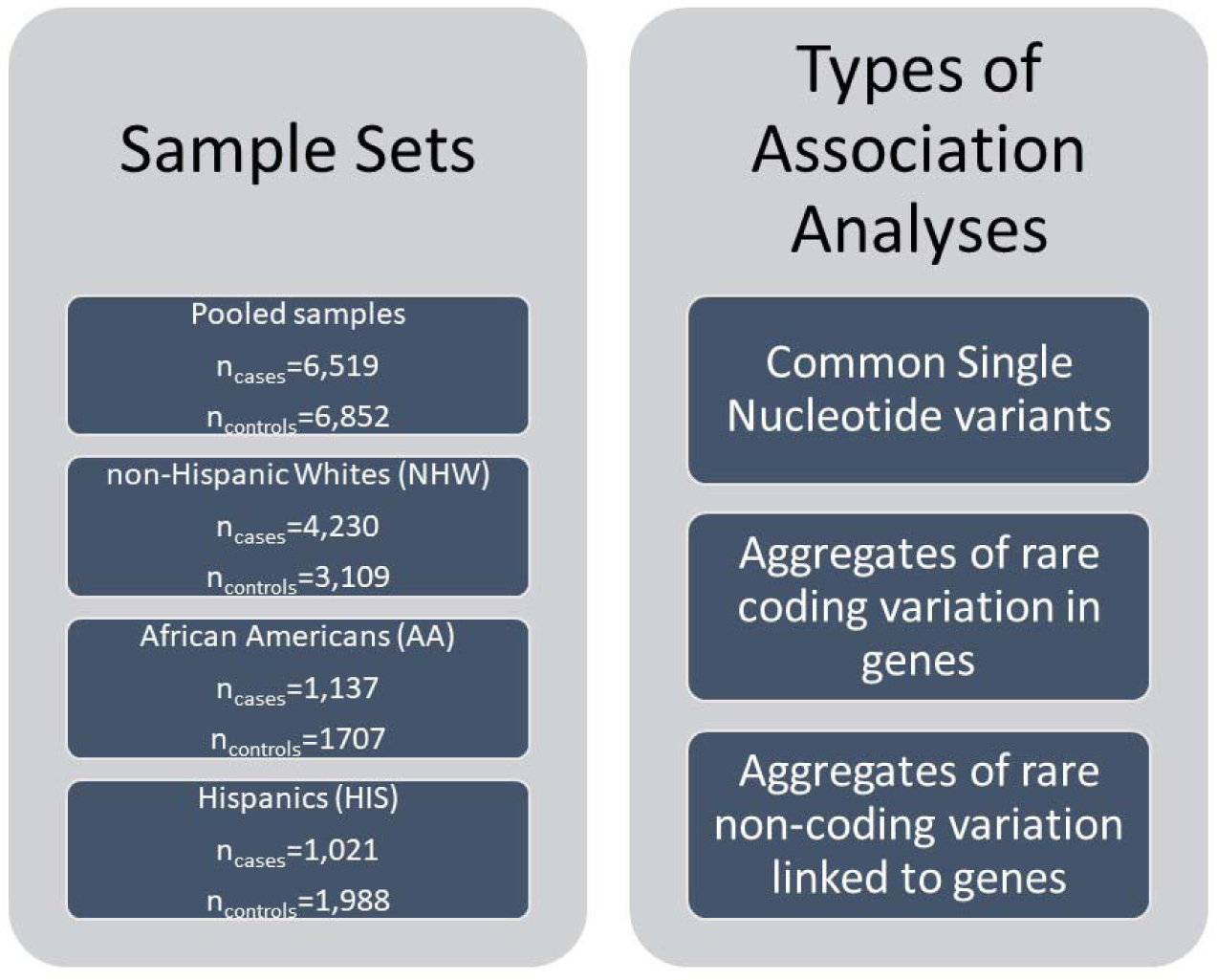
Study Overview. Three types of association analyses in four sets of individuals were performed, pooled samples, non-Hispanic Whites (NHW), African Americans (AA), and Hispanics (HIS). The pooled samples set included all individuals in the NHW, AA, and HIS sets, plus individuals that were not defined to be in one of those subsets.

**Table 1.** Descriptive statistics of ADSP study samples used for analyses.

|  | **AD**<br>**(N=6519)** | **No AD**<br>**(N=6852)** |
| --- | --- | --- |
| **Sex** |  |  |
| Female | 3921 (60.1%) | 4580 (66.8%) |
| **Age** |  |  |
| Mean (SD) | 75 (10) | 77 (8.0) |
| **APOE ε4 alleles\*** |  |  |
| 0 | 3022 (46.4%) | 4608 (67.3%) |
| 1 | 2917 (44.7%) | 1993 (29.1%) |
| 2 | 570 (8.7%) | 164 (2.4%) |
| **APOE ε2 alleles\*** |  |  |
| 0 | 6068 (93.1%) | 5930 (86.5%) |
| 1 | 427 (6.6%) | 800 (11.7%) |
| 2 | 14 (0.2%) | 35 (0.5%) |
| **Reported Race/Ethnicity** |  |  |
| non-Hispanic White (NHW) | 4230 (64.9%) | 3109 (45.4%) |
| Hispanic (HIS) | 1021 (15.7%) | 1988 (29.0%) |
| African American (AA) | 1137 (17.4%) | 1707 (24.9%) |
| Other | 131 (2.0%) | 48 (0.7%) |
| * ε2 and ε4 are derived from APOE genotype not WGS. | | |

### Association of Single Variants with AD

As expected, we found the strongest associations at the *APOE* locus (chr19:44,905,796-44,909,393), the major genetic risk factor for AD^8,24^. These associations were observed in the pooled samples analysis as well as the analysis within each of the three subgroups (**Figures 2 and S1**). We observed that the ⍰4 haplotype (the alternative allele at rs429358 and reference allele at rs4712) is more common in AD cases as well as most frequent in AA individuals and least frequent in HIS individuals (**Table S2**). While the odds of AD were higher in the AA individuals, the 95% confidence interval overlaps with the confidence interval for the NHW (**Figure 2**). We observed the ⍰2 haplotype (the reference allele at rs429358 and alternative allele at rs4712) was enriched in controls and had a lower frequency (frequency=0.05) than the ⍰4 haplotype (frequency=0.24). The ⍰2 haplotype is most frequent in the AA individuals, followed by HIS individuals, and least frequent in NHW individuals.

**Figure 2.**
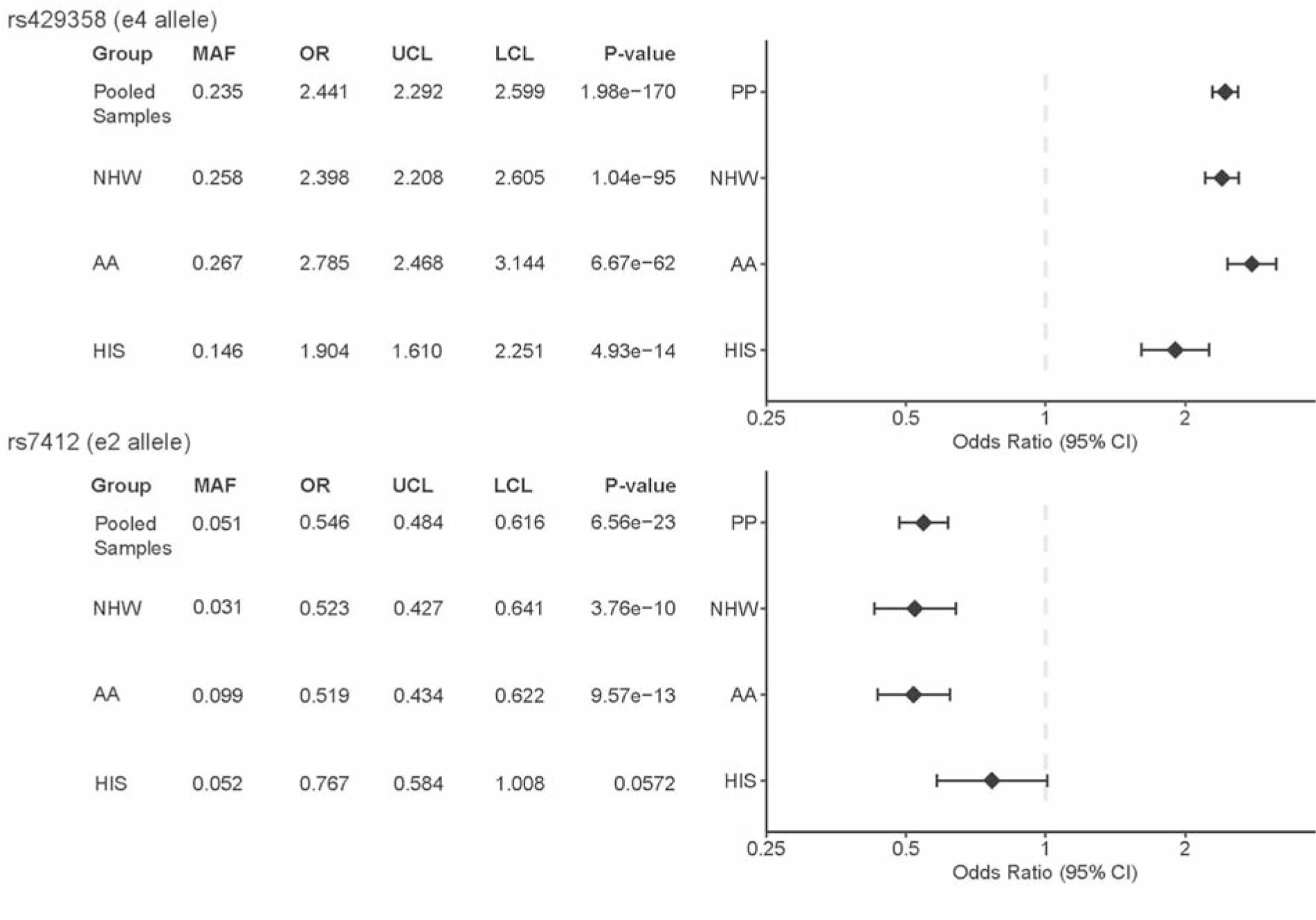
Association of APOE alleles with AD by subgroup. NHW, non-Hispanic White; AA, African American; HIS, Hispanic; OR, odds ratio; UCL, upper confidence limit; LCL, lower confidence limit

We also observed a genome-wide (GW) significant (p < 5×10^-8^) association of *BIN1* (rs4663105, MAF=0.47, OR=1.17, p=3.2×10^-9^) with AD status in the pooled samples analysis. Previous studies have identified *BIN1* as an AD susceptibility gene after *APOE*^12,14,25^. After adjusting for *APOE* ⍰4 (rs429358) and ⍰2 (rs7412) alleles on chromosome 19, the association of *BIN1* on chromosome 2 remained largely the same (OR=1.17, p=3.1×10^-9^). *BIN1* variants were also associated with AD in the NHW subgroup (MAF=0.43, OR= 1.22, p=1.2×10^-8^, **Tables 2** and **S3**). Although the association did not reach statistical significance, the *BIN1* variant showed a consistent direction of effect in the AA and HIS subgroups (AA: MAF=0.41, OR=1.16, p=0.0064; HIS: MAF=0.40, OR=1.10, p=0.15).

**Table 2.**
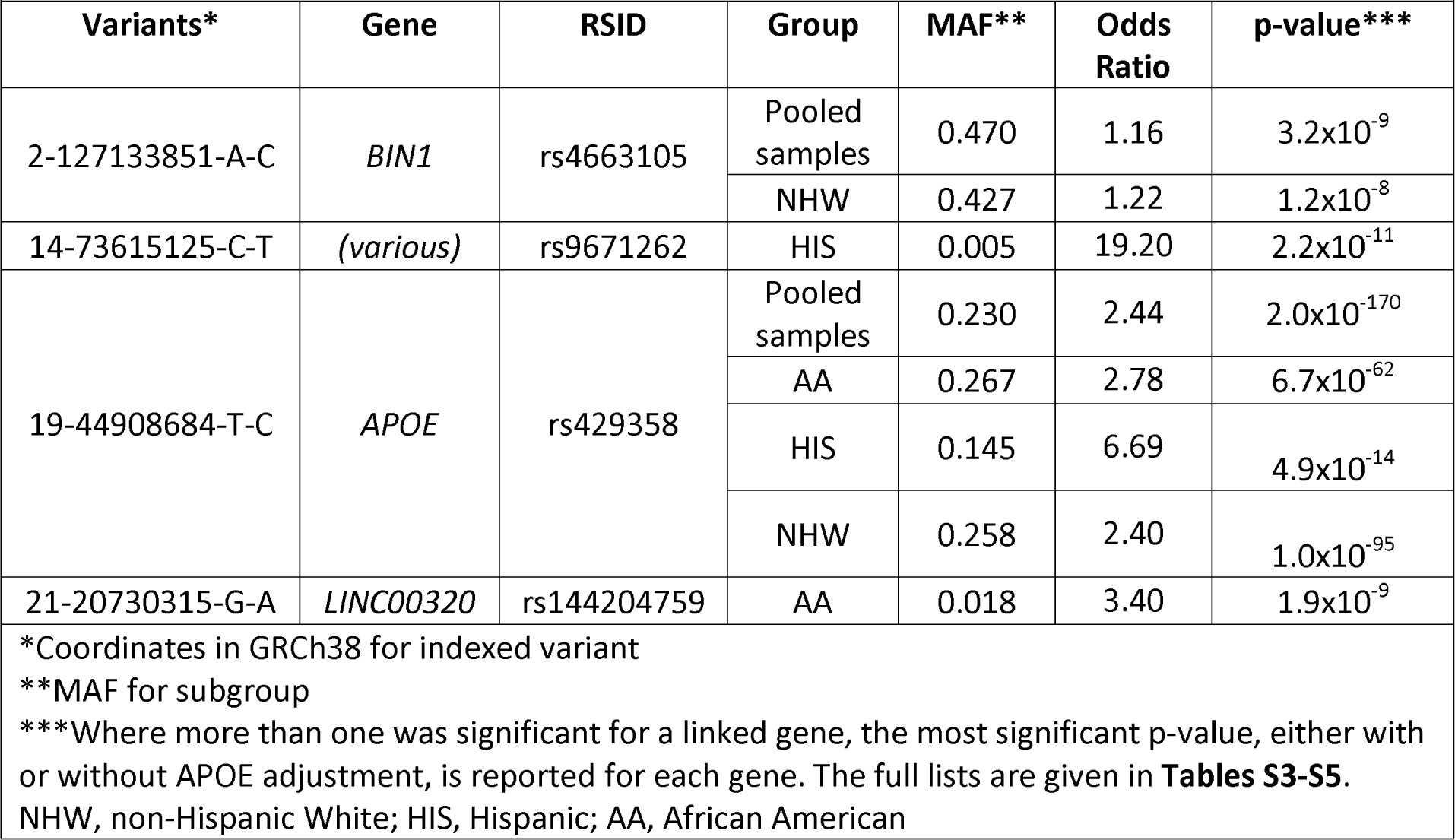
Genome-wide (p < 5×10^-8^) significant loci associated with AD.

In AA individuals, we observed an association between variants in *LINC00320* with AD (rs144204759, MAF=0.018, OR=3.4, p=1.9×10^-8^; **Tables 2** and **S4**, and **Figure S2A)**. This gene was previously implicated in AD^26^ by an distinct variant in the International Genomics of Alzheimer’s Project (IGAP) genome-wide association study (GWAS) of 25,170 AD cases and 41,052 cognitively normal controls. Additionally, three INDELs near *APOE* are associated with AD in the AA analysis (rs142042446, MAF= 0.040, OR= 2.27, p=5.7×10^-9^; rs542555887, MAF= 0.12, OR= 1.77, p=2.0×10^-10^; rs113492558, MAF=0.07, OR= 2.56, p=1.7×10^-19^); however, the reduction of these signals after conditioning *APOE ε2* and *ε4* alleles suggests that the impact of these INDELs is not distinct of these common *APOE* haplotypes.

### 14q24 in Hispanics

In Hispanic individuals, we observed a region from 14q24.2 to 14q24.3 (chr14:72,600,928-75,846,454), with 44 low frequency variants (43 SNVs and one INDEL; MAF: 0.005-0.012; **Tables 2** and **S5**) associated with AD across 13 genes (**Figures S1** and **S2**). These variants are not associated with AD in the AA subgroup despite a notably higher allele frequency (MAF: 0.007-0.041) (**Table S6**). Single variant analyses were not conducted for these variants in the pooled samples and NHW subgroups because the MAF in these subgroups was below the 0.5% threshold (**Table S6**). This region contains *PSEN1* p.G206A (rs63750082 at 14:73192712), a known early onset AD causal mutation^27–30^. *PSEN1* p.G206A was first identified in a few Caribbean Hispanic families^27,28^, and a follow-up study identified p.G206A in 70 families of Caribbean Hispanic ancestry^29^. A more recent study of early-onset AD in Hispanics in Florida found that 13 out of 27 participants (48.1%) were p.G206A carriers^30^. The allele frequency of this mutation is ultra-rare, only 1 in 1,741 to 3,790 individuals carries a *PSEN1* p.G206A in the Trans-Omics for Precision Medicine (TOPMed) Freeze 8^31^ and Genome Aggregation Databases (gnomAD) v3.1.2^32^ data, respectively. And while it is still ultra-rare, it is more common in Latino/Admixed Americans (1 in 423) within gnomAD v3.1.2. We observed 17 *PSEN1* p.G206A carriers in the entire ADSP R3 dataset (1 in 786), of which 16 individuals have AD, and all p.G206A carriers are reported as Hispanic. Only one pair of *PSEN1* p.G206A carriers were inferred to be related (**Figure S3**). Among those with AD, we observed that *PSEN1* p.G206A carriers have an earlier age of diagnosis of AD (58.6 +/− 7.6 years old) compared to non-carriers (74.7 +/− 10.4 years old; p=1.1×10^-10^).

Although previous studies have suggested that *PSEN1* p.G206A originated in Caribbean Hispanics, we do not have detailed data on geographic location of most ADSP participants. We applied principal components analysis (PCA) to decipher the origin of the p.G206A allele. PCA captures human population structure associated with ancestry^33^. Therefore, we placed p.G206A carriers into a global context using PCA of carriers and reference samples in the 1000 Genomes Project^34^ from 26 populations, including 139 Puerto Ricans. Expectedly, the 17 p.G206A carriers are closer to Puerto Rican reference samples in the American group according to the Euclidean distance of the first five principal components (**Figure S4**); individuals with at least 21 of 43 haplotype-associated SNVs are similarly placed near Puerto Ricans in PC space with greater dispersion (**Figure S5**). A global ancestry analysis revealed that all p.G206A carriers have a higher proportion of European (73.82% +/− 6.85%) than African ancestry (16.05% +/− 5.72%). However, we observed that the chr14 risk haplotype including p.G206A is inherited on an African-derived haplotype (**Figure S6**). As p.G206A is not detected in any individual of the AA subgroup while the other chr14 haplotype-defining variants have higher allele frequencies in the AA subgroup; p.G206A may be too rare to be detected on AA subgroup or may have arisen in Puerto Rico^35^ locally based on a founder event of an African haplotype. One female p.G206A carrier, whose *APOE* genotype is ε3/ε3 and global ancestry is 76.94% European genomes, was not diagnosed as an AD patient at the age of 74 years, 15 years older than the average age-at-onset of AD among p.G206A carriers. This raises the question of whether any other protective variants countervail the impact of mutations in the haplotype. To the best of our knowledge, this is the first report of a significant association between AD and this 3Mbp haplotype, which was linked to onset age 20 years younger^27,28^.

### Association of Aggregates of Rare Variants in Genes

We performed gene-centric aggregates of coding variants for each protein-coding gene using 5 functional variant categories (putative loss of function (pLoF), missense, disruptive missense, pLoF and disruptive missense, and synonymous). Unsurprisingly, we observed several significant gene-based associations (p<5×10^-7^) in 14q24 in the HIS individuals (**Table S7**). Additionally, we observed that *PSEN1* was associated with AD (p = 4.1×10^-8^) in the pooled samples analysis when aggregating rare loss of function and disruptive missense variants (**Figure 3**). Sensitivity analyses excluding p.G206A (rs63750082) from the gene-based tests revealed a significant association between the aggregation of rare loss-of-function and disruptive missense variants in *PSEN1* and AD in the pooled samples individuals (OR 2.02, 95% CI 1.3-3.0, p=7.8×10^-4^). These results suggest that there may be additional rare variants in *PSEN1* that contribute to the observed association with AD, particularly in the NHW subset.

**Figure 3.**
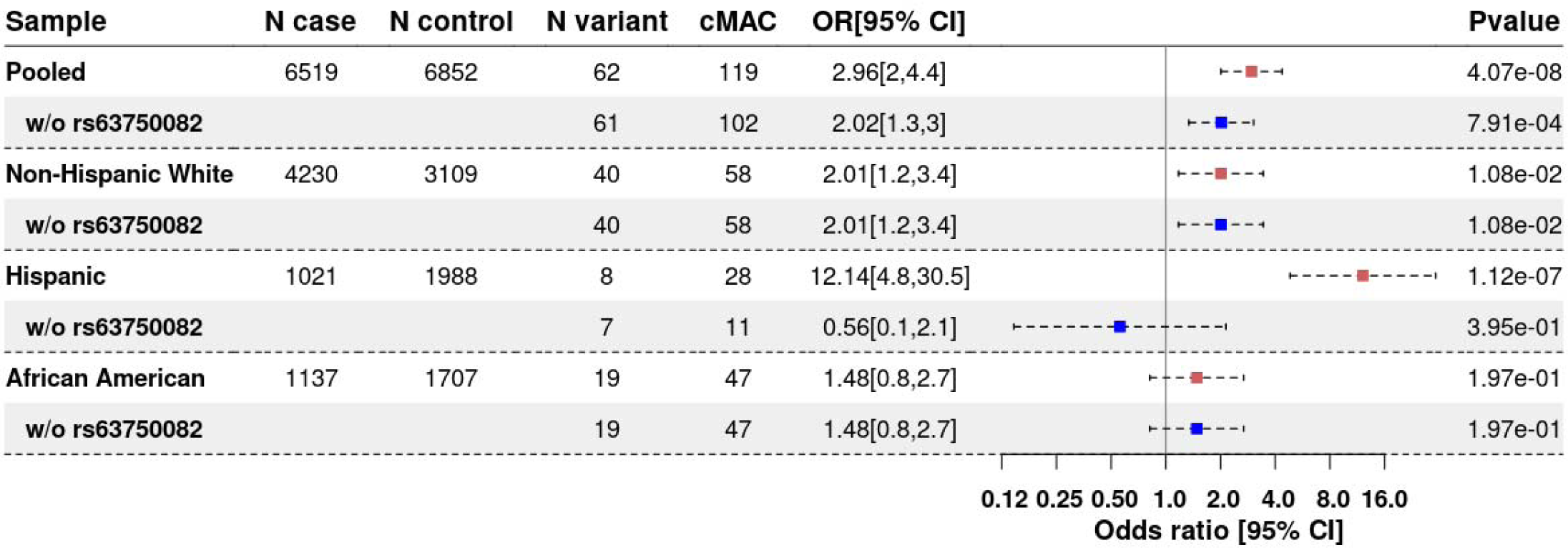
Association of *PSEN1* with AD by subgroup, with and without G206A included.

We observed a few suggestive gene-based associations driven by rare coding variants (5×10^-7^ < p < 5×10^-5^). Significant association signals between AD and *ABCA7* (p = 5.4×10^-6^) in the NHW analysis and *SPTLC2* (p = 1.0×10^-5^) in the HIS analysis were observed using multiple sets of rare coding variant aggregates (i.e., loss of function plus disruptive missense as well as missense, **Table S7**). There has been a growing interest in studying *ABCA7* due to accumulating *in vitro* and *in vivo* studies supporting the potential contribution to AD-related phenotypes^36,37^. In the NHW subgroup, there are 273 rare exonic variants in *ABCA7* (including 186 nonsynonymous, 9 stop-gain, 7 frameshift, and 3 non-frameshift; **Table S8**), and 90 of them are not singletons (including 67 nonsynonymous, 2 stop-gain and 4 frameshift; MAF: 0.000068 - 0.0075). Two frameshift deletions, rs547447016 (chr19:1047508:AGGAGCAG:A; N_case_=30 and N_control_=9) and rs538591288 (chr19:1055908:CT:C; N =20 and N =8), have been reported in previous studies^38–40^ and experimentally validated. Another two, rs779501556 (chr19:1046404:CGT:C; N_case_=2 and N_control_=0) and rs745871063 (chr19:1054250:AG:A; N_case_=0 and N_control_=2), are novel and ultra-rare. Nine of the stop-gain SNVs in our *ABCA7* test (**Table S8**) were previously identified associated with AD^38,41–45^ or Autism^46^.

*SPTLC2* (chr14:77,505,997-77,616,637) encodes a protein involved in the biosynthesis of sphingolipids, and there is some evidence to suggest that changes in sphingolipid metabolism may be associated with AD^47,48^. Specifically, previous studies have suggested that alterations in the levels of specific sphingolipids may contribute to the development or progression of the disease^47,49^. However, the role of *SPTLC2* in AD is not yet fully understood, and research in this area is ongoing. Our WGS study identified a suggestive significant association between the aggregate of rare disruptive missense and loss of function variants in *SPTLC2* (**Table S9**) with AD in the HIS subgroup (p = 1.0×10^-5^ and 1.0×10^-5^), respectively.

### Association of Aggregates of Rare Variants in Noncoding Sets

We next performed gene-centric aggregates of rare noncoding variants using 8 functional variant categories (promoter or enhancer overlaid with CAGE or DHS sites, UTR, upstream, downstream, and noncoding RNA genes). We observed rare noncoding variant aggregates associated with AD near *TOMM40* (p=7.2×10^-8^) and *PSEN1* (p=2.4×10^-11^ to 3.2×10^-8^) regions in the pooled samples and HIS individuals (**Table 3**). After conditioning on the number of *APOE* ε2 and ε4 alleles, there was an attenuation of the results for *TOMM40* in the pooled samples analysis, but the association remained (p <7.1×10^-6^). Compared to the pooled samples analysis, we observed that none of subgroup-specific associations reached a GW significant level for our rare variant aggregation tests (p<1×10^-7^), however, we observed suggestive associations signals in the AA and NHW individuals with variants in the promoter of *TOMM40*, p=2.8×10^-4^ (p_adj_=0.010) and 4.4×10^-3^ (p_adj_=0.018), respectively, whereas there was not an association in HIS individuals (p=0.41, p_adj_=0.45). After adjusting for *PSEN1* p.G206A, we observed that the association of rare noncoding variants in the promoter of *PSEN1* was no longer significant (p>0.05) in pooled samples and HIS analyses. This suggests these rare noncoding variants are on the same haplotype as the rare coding variants in *PSEN1*; this was confirmed by local ancestry analysis (**Figure S6**).

**Table 3.** Aggregates of rare variants in noncoding sets with AD.

| **Group\*\*** | **Gene name** | **Chr** | **Category** | **# variants** | **STAAR-O p-value\*** |
| --- | --- | --- | --- | --- | --- |
| Pooled samples | *TOMM40* | 19 | Promoter (DHS) | 134 | $7.2 \times 10^{-8}$ |
| Pooled samples | *ELMSAN1* | 14 | Enhancer (DHS) | 1133 | $1.8 \times 10^{-9}$ |
| Pooled samples | *EIF2B2* | 14 | Enhancer (DHS) | 1240 | $3.2 \times 10^{-8}$ |
| Pooled samples | *MIR4505* | 14 | ncRNA | 7 | $2.4 \times 10^{-11}$ |
| HIS | *PTGR2* | 14 | Promoter (CAGE and DHS) | 7 | $8.85 \times 10^{-12}$ |
| HIS | *ELMSAN1* | 14 | Enhancer (DHS) | 366 | $3.1 \times 10^{-11}$ |
| HIS | *PTGR2* | 14 | Enhancer (CAGE) | 153 | $5.9 \times 10^{-11}$ |
| HIS | *ACOT6* | 14 | Enhancer (DHS) | 33 | $4.2 \times 10^{-10}$ |
| HIS | *ELMSAN1* | 14 | Promoter (DHS) | 55 | $8.8 \times 10^{-10}$ |
| HIS | *ACOT4* | 14 | Promoter (CAGE) | 6 | $9.3 \times 10^{-10}$ |
| HIS | *ACOT4* | 14 | Enhancer (CAGE) | 9 | $9.8 \times 10^{-10}$ |

\*Where more than one rare noncoding aggregate was significant for a linked gene, the most significant p-value is reported for each category type.

### Spurious Variants

Six SNVs in *ANK3* were associated with AD from the NHW single variant analysis (**Table S10**). However, after a closer investigation, all these variants were false positive associations. These variants were initially considered to be of high quality with reasonable ABHet (allele balance at heterozygous sites) values (0.69-0.72) and passed the Hardy-Weinberg Equilibrium (HWE) test by RUTH^50^. However, we noted that all six alternate alleles were almost always on the same sequencing read. Inspection of these data revealed that these variants were supported only by supplementary or improperly-paired reads with mapping quality <= 6. We therefore excluded these variants from our analysis given their poor support. After checking alignments, it was determined that these variants were from supplementary or improper-paired alignments with mapping quality ≤ 6 (**Figure S7**). Due to the lack of certainty regarding alignment quality, these variants were filtered out.

Our analysis identified 12 INDELs (**Table S10**) associated with AD from the single variant analysis, but none were confirmed by experimental validation. Further investigation revealed most of them are located in poly-A regions or discrepancies between the sequences in the regions of the genome from Telomere-to-Telomere Consortium (T2T)^51^ and GRCh38 (the reference genome we used), indicating a potential bias issue with the reference genome. GRCh38 is constructed from a single haplotype and may not accurately represent the genomic diversity of humans, leading to incorrect mappings of short reads from a sample and resulting in false positive variant calls. Our findings highlight the need for best practices in handling INDELs to avoid potential biases and improve the accuracy of genetic variant calls.

## Discussion

WGS data allow for the testing of both common and rare genetic variation that may be unique to individuals or populations and provide a powerful tool for identifying genetic variation that may be missed by traditional genotyping methods. Since the ADSP includes the largest sample of diverse participants with WGS and AD status, we designed our study to perform association testing across all participants and within subgroups defined by reported race/ethnicity. The pooled samples analysis is most powerful to detect associations when there are similar effects across ancestries, while the subgroup-specific analyses are able to detect subgroup-specific effects. Through our analysis framework, we were able to adequately control for the diversity within ADSP and leveraged ADSP WGS to learn more about known loci for AD, including population-specific genetic signals.

As anticipated, our single variant GWAS showed the strongest associations with AD in *APOE* across all groups as well as *BIN1* for the pooled samples and NHW. We observed that genetic variation in *LINC00320* was associated with AD in AA. We identified 44 genetic variants on 14q24 in HIS (MAF: 0.005-0.012), and aggregates of rare variants analysis in HIS and pooled samples confirmed the region, which includes p.G206A in *PSEN1*, a well-known early onset AD mutation^27–30^. Our PCA analysis indicates that p.G206A carriers are closer to Puerto Ricans, consistent with previous studies^35^, while local ancestry analysis, however, pointed out that the local haplotype is derived from African genomes.

The analysis of coding rare variants identified suggestive associations in *ABCA7* in NHW and *SPTLC2* in HIS with AD. Rare coding variants in *ABCA7* have been associated with AD risk in AA individuals^52,53^, however, we additionally observe an association in the NHW. A deeper investigation of *ABCA7* revealed two frameshift deletions, rs547447016 and rs538591288, that were reported in previous studies^38–40^ and validated here, while rs779501556 and rs745871063 are variants newly associated with AD in the current analysis, indicating distinct *ABCA7* variants in multiple subgroups associated with AD risk. The noncoding rare variants in the promoter of *TOMM40* were identified as significant in the pooled samples analysis and confirmed to be distinct from the *APOE* haplotypes.

There are some limitations to our study. First, defining subgroups based on reported race and ethnicity, which was used in this study, may not be recommended as the best practice^54^, we observed a high consistency in reported and genetically-defined subgroups, particularly for the AA and NHW subgroups (**Table S11**). Furthermore, humans cannot be grouped into discrete categories, and we acknowledge heterogeneity in our subgroup-specific analyses, and that the heterogeneity varies among the subgroups. We clustered samples by using the Euclidean distance of PCA between each individual and the three 1000 Genomes Project reference populations, Europeans (EUR), Africans (AFR), and East Asians (EAS) (**Figure S8**). Three subgroups were formed, e.g., AA-AFR (AA samples closer to AFR), HIS-EUR (HIS samples closer to EUR), and NHW-EUR (NHW samples closer to EUR). Association analyses on genetic-defined subgroups gave similar results. Furthermore, from our global ancestry analysis, 94.13% reported African Americans (AA) are inferred as having a majority of African ancestry, and 99.43 reported Non-Hispanic Whites (NHW) are inferred as having a majority of European ancestry. As to the reported Hispanics (HIS), 74.74%, 20.51%, 2.43%, and 1.46% are inferred by GRAF-pop, a global ancestry inference, as having ancestries of Latin American 1, African American, Latin American 2, and European, respectively from GRAF-pop^55^. Secondly, despite being the largest sample with WGS ascertained on AD status, we still have limited power to detect associations with AD, particularly with rare noncoding variants where the aggregation unit is not as well defined as with rare coding variants. We acknowledge there are much larger sample sizes using GWAS for AD to assess the contribution of single variants^12^.

In conclusion, we have comprehensively analyzed up to 13,371 diverse individuals with WGS for AD and observed common and rare variants associated with AD.

## Methods

### Study Participants

Data from the ADSP are available to qualified investigators via the National Institute on Aging Genetics of Alzheimer’s Disease Data Storage Site (NIAGADS) (https://dss.niagads.org/). The current analyses focused on participants with WGS in the NIAGADS data named “R3 17K WGS Project Level VCF”. WGS data have been generated in multiple cohorts as part of the ADSP. The Release 3 (R3) includes whole-genome data from 1,020 ADSP Family Discovery and Discovery Extension samples, 2,959 ADSP Case Control Extension samples, 809 ADNI-WGS-1 samples, 886 CurePSP and Tau Consortium PSP samples, 408 PSP UCLA samples, 617 NINDS, CurePSP and Tau Consortium PSP samples, 209 University of Pittsburgh-Kamboh samples, 207 Cache County samples, 77 Knight ADRC samples, 91 FASe_families samples, 137 NACC-Genentech samples, 730 AMP-AD ROSMAP samples, 344 AMP-AD MSSM samples, 252 AMP-AD MAYO samples, and 8,160 ADSP Follow-Up Study 1 samples (FUS1 contains 885 ADSP FUS1 APOE Extremes samples, 2,771 ADSP FUS1 ADC Autopsy samples, 1,517 ADSP FUS1 PR1066 samples, 1,923 ADSP FUS1 ADGCAA samples, 757 ADSP FUS1 ADNI-WGS-2 samples, 92 ADSP FUS1 Miami HIHG Brain Bank samples, and 214 ADSP FUS1 StEP-AD samples). The Discovery phase WGS was generated for individuals from multiplex AD families as previously described^23,56,57^. The Discovery Extension phase consisted of a family component and a case control component. The Discovery Extension family component WGS was generated on additional members of selected families from the Discovery phase as well as members of 77 additional families. A set of 114 Hispanic control individuals were also sequenced with the family component. A focus of the Discovery Extension case control component was to increase the diversity of the ADSP samples. In the ADSP Discovery and Discovery Extension phases sequencing was performed at three sequencing centers via the National Human Genome Research Institute (NHGRI). Sequence data for ADSP Augmentation Studies were generated under NIA and private funding and are shared with the research community via NIAGADS. The ADSP Follow-up Study (FUS) (https://adsp.niagads.org/the-alzheimers-disease-sequencing-project-adsp-follow-up-study-fus/) contains individuals with existing cognitive data with the ability to adjudicate Alzheimer disease status with whole genome sequencing performed at the American Genome Center at the Uniformed Services University of the Health Sciences (USUHS) in coordination with existing NIH-funded AD infrastructure including the National Cell Repository for Alzheimer’s Disease (NCRAD), NIAGADS, and the Genome Center for Alzheimer’s Disease (GCAD). The ADSP data coordinating center, the Genomic Center for AD (GCAD), produced a jointly called and quality controlled (QC’ed) data set for WGS. Details of studies included in the ADSP can be found at NIAGADS under dataset: NG00067 ADSP Umbrella Study (https://dss.niagads.org/datasets/ng00067/). Of the 16,905 individuals in the ADSP Release 3 data, individuals with unknown AD status and genetically identical individuals were excluded. After removing these individuals, 13,371 individuals were available for association analysis with AD.

We created three groups for subgroup-specific analyses. Individuals who reported their race as White and their ethnicity as Non-Hispanic or missing were classified as non-Hispanic White (NHW). Individuals who indicated any reported race and Hispanic ethnicity were classified as Hispanic (HIS). Individuals who reported their ethnicity as Non-Hispanic or had missing ethnicity and their race as Black were classified as African American (AA). There were 179 individuals that were not classified into one of our subgroup-specific analyses. Using genetic similarity clustering did not substantially change the subgroups.

### Sample Clustering Using Genetic Similarity

To cluster samples based on genetic information, we employed the following approach. Firstly, we calculated the Euclidean distance of PC1-2 between each sample and the three reference populations from the 1000 Genomes Project, Europeans (EUR), Africans (AFR), and East Asians (EAS) (**Figure S8**). The results indicated that 92.94% and 7.06% of reported African American (AA) samples were found to be closer to the AFR and EUR reference populations, respectively. Additionally, 8.28%, 0.003%, and 91.69% of reported Hispanic (HIS) samples were closer to the AFR, EAS, and EUR reference populations, respectively. Similarly, 0.08%, 0.01%, and 99.90% of reported non-Hispanic white (NHW) samples were closer to the AFR, EAS, and EUR reference populations, respectively. Based on these findings, we clustered the samples into subgroups: AA-AFR (AA samples closer to AFR), HIS-EUR (HIS samples closer to EUR), and NHW-EUR (NHW samples closer to EUR). Subsequently, we performed association analyses on these PC defined subgroups. However, the results of these analyses did not show significant changes compared to the previous sub-group analyses based on reported race and ethnicity. We employed a second approach to create sample clusters, which involved conducting global ancestry inference analysis. This analysis revealed that 94.13% of reported AA samples were inferred to have a predominantly African ancestry, while 99.43% of reported NHW samples were inferred to have a predominantly European ancestry. Regarding reported HIS samples, 74.74% were inferred to have Latin American 1 ancestry, 20.51% had African American ancestry, 2.43% had Latin American 2 ancestry, and 1.46% had European ancestry.

### AD Phenotype Definition

The ADSP provides different AD status variable definitions for participants included via case-control versus family-based studies. Additionally, distinct phenotype data are available for some augmentation studies. In the current analysis, for individuals in the ADSP case-control study, we defined AD cases as individuals with either prevalent or incident AD. Case-control individuals with no prevalent or incident AD were defined as controls and those with NA for status were defined as unknown. In the ADSP family phenotype file, possible values for the AD status variable include no dementia, definite AD, probable AD, possible AD, family-reported AD, other dementia, family reported no dementia, and unknown. For family-based individuals, we defined an AD case as either possible, probable or definite AD. AD controls were defined as individuals coded as no dementia. We redefined individuals with family-reported AD, other dementia, or unknown status as missing AD status. The ADNI phenotype data, which is part of the ADSP augmentation study, provides information on mild cognitive impairment (MCI) in addition to AD status. Individuals with a current diagnosis of MCI were included as AD controls in the current study.

### Principal Component Analysis

Principal Component Analysis (PCA) is widely used for analyzing large datasets that have a high number of dimensions or features per observation. PCA is a statistical technique for reducing the dimensionality of a dataset, while still retaining as much of the original variation as possible. In genetic studies, PCA is commonly used to infer population structure in the data, since population structure is a major factor that affects sample genotypes. Typically, the top principal components (PCs) calculated from the genotype data reflect population structure among the individuals. To ensure accurate ancestry inference, we used PC-AiR in the GENESIS^58^ package for a PCA, a tool that accounts for sample relatedness and thus provides accurate ancestry inference. We calculated PCs for the pooled samples using variants with MAF > 5%, call rate > 99%, GCAD provide variant flag (VFlag)=0 (no exclusion), Ruth HWE p-value > 10^-4^ and excluded variants in high LD regions (https://genome.sph.umich.edu/wiki/Regions_of_high_linkage_disequilibrium_(LD)) and variants in LD using an r2 < 0.1,. For the AA, HIS, and NHW subgroups, we calculated PCs using SNVs with MAF > 5%, VFlag^23^ = 0, and LD threshold r2 (0.1).

To identify the locations of G206A carriers, we extended the PC calculation to include samples of the 1000 Genomes Project. The VCFs of the 1000 Genomes Project were downloaded and merged with ADSP R3 VCFs by bcftools. Population and super-population information of each sample was also downloaded from the 1000 Genomes Project. The MAF cutoff, >5%, was applied, and then PC calculation was performed.

### Covariates, Analysis Models, and Single Variant Analysis

We included sex, technical sequencing variables (sequencing center and sequencing length), and principal components (PC1-5 for subgroup-specific analysis and PCs associated with AD status for the pooled samples analysis) as covariates in all our models along with a genetic relationship matrix to adjust for relatedness among individuals. As a secondary model, we additionally adjusted for the number of APOE *e4* alleles and number of APOE *e2* alleles. We tested each variant with a MAF > 0.5% for association with AD using the score test in the GENESIS^47^ package to fit a penalized quasi-likelihood (PQL) approximation to the generalized linear mixed model. Variants that failed Hardy-Weinberg Equilibrium (HWE) by RUTH^42^ (HWE_SLP_I < −4 or HWE_SLP_I > 4) in controls were excluded as well as variants in low complexity regions. We used a standard significance threshold of 5×10^-8^ for our single variant association analyses.

### Aggregates of Rare Variants Analysis

To test aggregates of coding and noncoding rare variants, we implemented the STAAR pipeline^59^ using both SNVs and INDELs. The STAAR pipeline is a set of routines for performing association analysis of large-scale WGS data using the STAAR framework^60^ to aggregate rare variants using variant set analysis for both gene-centric coding and gene-centric non-coding analysis. We used the STAAR-O p-value, which combines p-values across multiple annotation-weighted variant set tests^60^.

The gene-centric coding analysis of the STAAR pipeline provides five genetic categories: putative loss of function (pLoF), missense, disruptive missense, pLoF and disruptive missense, and synonymous. The gene-centric noncoding analysis provides eight genetic categories: promoter or enhancer overlaid with CAGE or DHS sites, UTR, upstream, downstream, and noncoding RNA genes. We set our significance threshold for our rare variant aggregation tests to be 1×10^-7^ (Bonferroni correction for testing ∼20,000 genes across 5 coding categories and 8 non-coding categories). For gene-centric noncoding analysis, due to the known associations in *PSEN1* and *APOE* regions, we performed conditional analyses adjusting for p.G206A (rs63750082) in chromosome 14 and *APOE e2* and *e4* alleles in chromosome 19 in the pooled samples analysis. To incorporate additional features of the STAAR pipeline, we created a github repository that performs variant extraction and conditional analysis (see code availability).

### Global and Local Ancestry Inference Analysis

The global ancestry inference was performed using the GRAF-pop^55^ tool, which utilizes 100,437 fingerprint SNPs and is a PCA-free method for ancestry inference. GRAF-pop provides results of comparable quality to PCA-based methods such as EIGENSTRAT, FastPCA, and FlashPCA2, while offering an ultra-fast running time. Genotypes were provided to GRAF-pop in VCF format. The tool assumes that each individual is a mixture of three ancestries: European (E), African (F), and Asian (A), and estimates the ancestral proportions *Pe*, *Pf*, and *Pa* using barycentric coordinates.

To infer local ancestry, specifically for the analysis in the 14q24 region, we utilized RFMIX^61^ with 2,504 reference genomes from the 1000 Genomes Project. This tool outperforms other methods for estimating local ancestry in complex admixture scenarios^62^. Prior to using RFMIX, we phased variants using SHAPEIT4^63^. In addition, we utilized PICARD (http://broadinstitute.github.io/picard/) to liftover coordinates from HG38 to HG19 as the genetic map of reference samples we used was against HG19 (https://mathgen.stats.ox.ac.uk/impute/1000GP_Phase3.html). RFMIX allows for a comprehensive understanding of the local ancestry composition in the specific region of interest and provides insights into the complex genetic makeup of diverse populations.

### INDEL Experimental Validation

PCR Primer Design: Genomic sequence for the INDEL variants was determined by submitting the chromosomal location of the variants to the Dec. 2013 (GRCh38/hg38) version of the Genome Browser^64^ (http://genome.ucsc.edu). Sequence surrounding the variants was extracted and used to design PCR primers. Primers were designed outside of the breakpoints to amplify across the insertion/deletion sequence. All PCR primer sequences were submitted to the Blast-like alignment tool (BLAT) to check for uniqueness of the sequences. When available, samples from three individuals reported as homozygous or heterozygous for the variant were used for sequence validation along with one control (or reference) sample. When possible, samples from multiple families were used for validation.

PCR and Sanger Sequencing: Genomic DNA (∼50ng) was amplified using a SimpliAmp Thermal Cycler (Applied Biosystems) in a 20ul reaction volume with HotStarTaq Master Mix (Qiagen) in the presence of 2uM primers (IDT). The PCR conditions used were: 95°C 15 min followed by 30 cycles of 95°C 20 sec, 55°C 30sec, 72°C 2min with a final extension of 72°C 7 min. The amplified PCR products were prepared for Sanger sequencing by adding ExoSAP-IT (USB) and incubating at 37°C for 45 min followed by 80°C for 15 min. The PCR products were then Sanger sequenced using the BigDye® Terminator v3.1 Cycle Sequencing kit (Part No. 4336917 Applied Biosystems). The sequencing reaction contained BigDye® Terminator v3.1 Ready Reaction Mix, 5X Sequencing Buffer, 5M Betaine solution (Part No. B0300 Sigma) and 0.64uM sequencing primer (IDT) in a total volume of 5ul. The sequencing reaction was performed in a SimpliAmp Thermal Cycler (Applied Biosystems) using the following program: 96°C 1 min followed by 25 cycles of 96°C 10 sec, 50°C 5 sec, 60°C 1min15sec. The products were cleaned using XTerminator and SAM Solution (Applied Biosystems) with 30 min of shaking at 1800 rpm followed by centrifugation at 1000 rpm for 2min. The sequencing products were analyzed on a SeqStudio Genetic Analyzer (Applied Biosystems) and the sequencing traces were analyzed using Sequencher 5.4 (Gene Code).

### INDEL in silico Validation

Based on our findings in this study, we encountered 12 significant INDELs that were experimentally validated as false positives. These INDELs were all situated within sequences that exhibited discrepancies between the Telomere-to-Telomere Consortium (T2T) ^43^ and GRCh38 (the reference genome utilized in our study). Consequently, INDELs located in these discrepant sequences between T2T and GRCh38 were not included in our report.

To address this issue, we initially identified regions by considering the coordinates of the INDELs along with their respective lengths, and then extended these regions by +/− 10 bp. Subsequently, we used liftover, using the hg38-to-chm13v2 chain (available at https://hgdownload.soe.ucsc.edu/goldenPath/hs1/liftOver/hg38-chm13v2.over.chain.gz) to convert the regions to the corresponding coordinates in the T2T (chm13v2) assembly. In cases where the liftover process was unsuccessful, the INDELs were excluded from further analysis. For the regions that successfully underwent liftover, we compared the sequences obtained from GRCh38 and T2T. Only when the sequences were found to be identical, we included the corresponding INDELs in our report, ensuring accuracy and reliability in our findings.

## Data availability

ADSP R3 VCFs: https://dss.niagads.org/datasets/ng00067/

1000 Genomes Project VCFs: http://ftp.1000genomes.ebi.ac.uk/vol1/ftp/data_collections/1000G_2504_high_coverage/working/20201028_3202_raw_GT_with_annot/20201028_CCDG_14151_B01_GRM_WGS_2020-08-05_chr$chr.recalibrated_variants.vcf.gz

## Code availability

https://github.com/wanpinglee/CADRE_CHARGE_ADSP17K https://github.com/seuchoi/STAARpipeline_plugin

## Acknowledgements

See supplementary text. WPL reports grant support from RF1-AG074328 and P30-AG072979. HL reports grant support from U01AG068221. WB, EM, and JLH reports grant support from U01 AG058654. JD, MF, EEB, ALD and GMP reports grant support from U01 AG058589. LAF reports grant support from U01-AG058654, R01-AG048927, U19-AG068753, U01-AG062602, U01-AG032984, U54-AG058654, P30-AG072978. MP-V reports grant support from U01 AG072547 and U01 AG070864. EMW reports grant support from U01AG058589 and P30AG066509. L-SW reports grant support from U24-AG041689, U54-AG052427, U01-AG032984, U01-AG058654, P30AG072979. XZ reports grant support from U01AG072577, R01AG080810.

## Author Contributions

WPL, SHC, MS, BAD, FN, and GMP performed statistical analyses. NLHC, AMB, LAF, MPV, LSW, JLH, GMP, and XZ performed phenotype acquisition and/or harmonization. WPL, ANP, AMB, YYL, HL, WSB, EM, EEB, XZ, MPV, LSW, ALD, JLH, and GMP performed Genotype acquisition and/or QC. WPL, SHC, P-LC, HW, HL, JD, LAF, EEB, XZ, MF, EM, EMW, LSW, ALD, JLH, and GMP interpretated results. WPL and GMP wrote the first draft of the manuscript. All authors read, critically revised, and approved the manuscript.

