## Supplementary Information for "Association of Common and Rare Variants with Alzheimer’s Disease in over 13,000 Diverse Individuals with Whole-Genome Sequencing from the Alzheimer’s Disease Sequencing Project"

### Supplementary Figures

#### Supplementary Figure 1. Manhattan and quantile-quantile QQ plots of the analyses prior to filtering false discovered SNPs.

(A) Pool-population (PP) analysis without APOE E2 and E4 adjustment

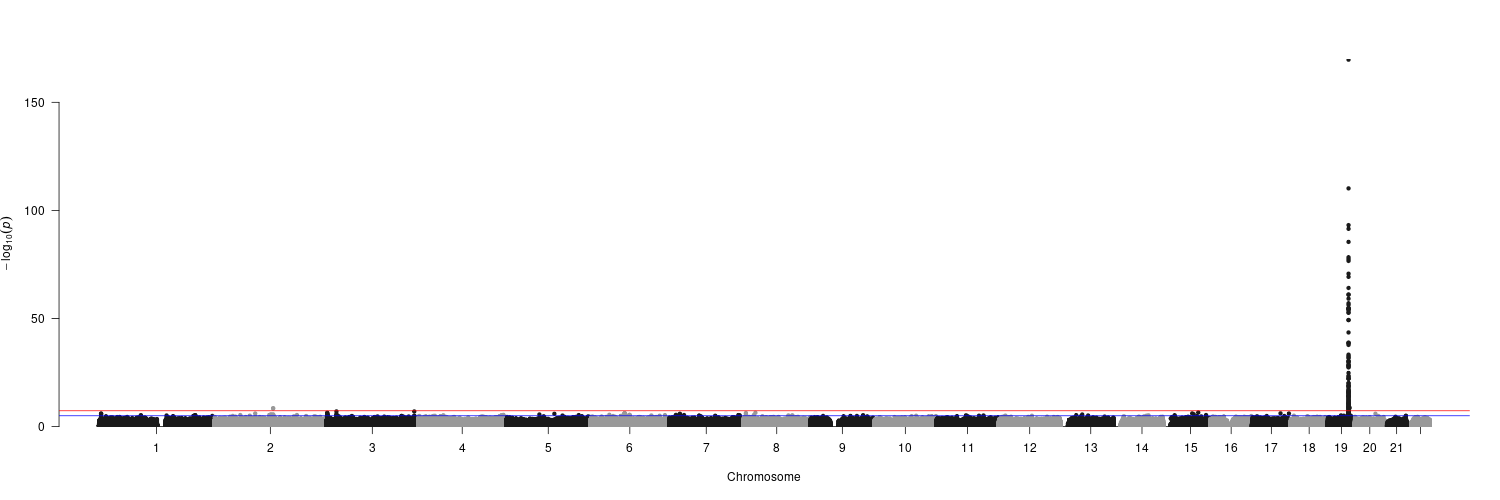

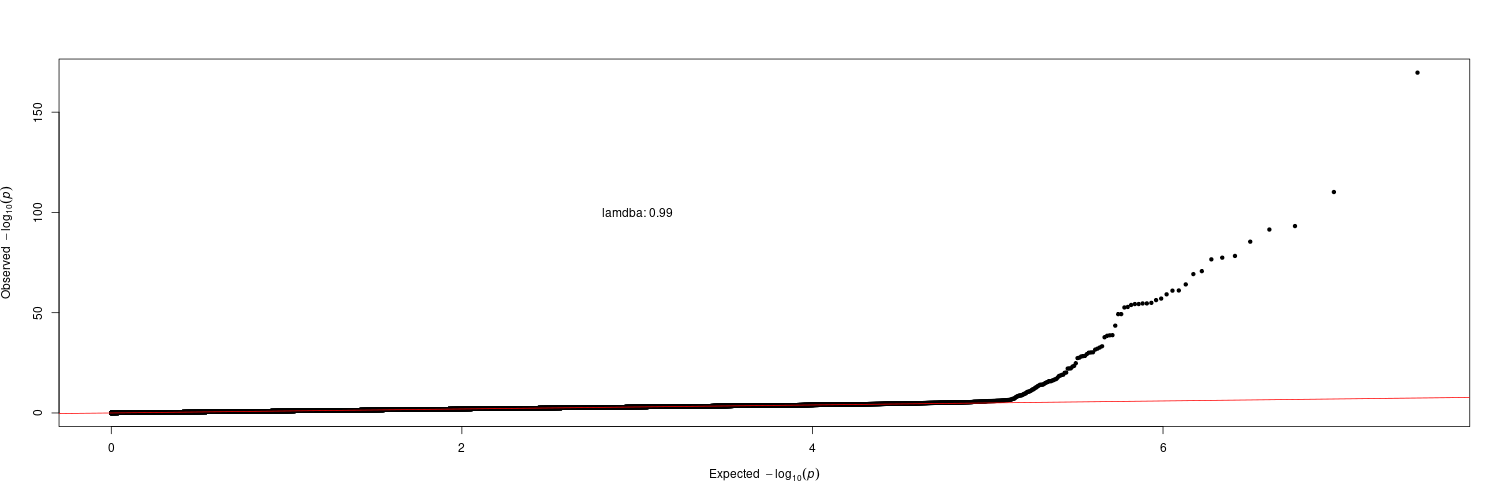

(B) Pool-population (PP) analysis with APOE E2 and E4 adjustment

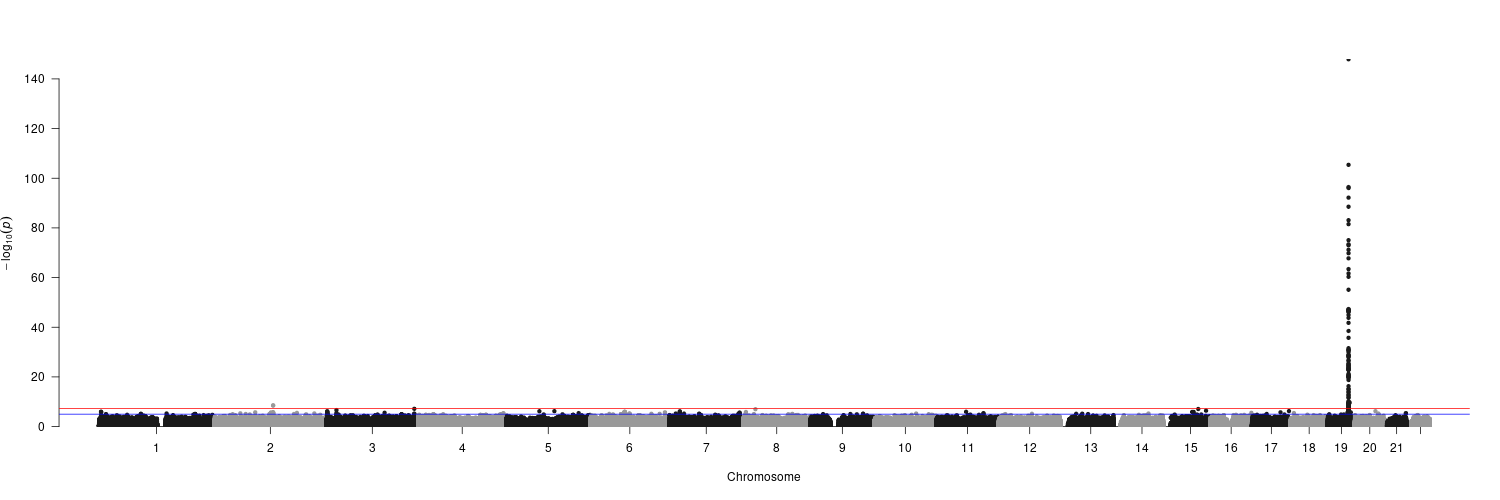

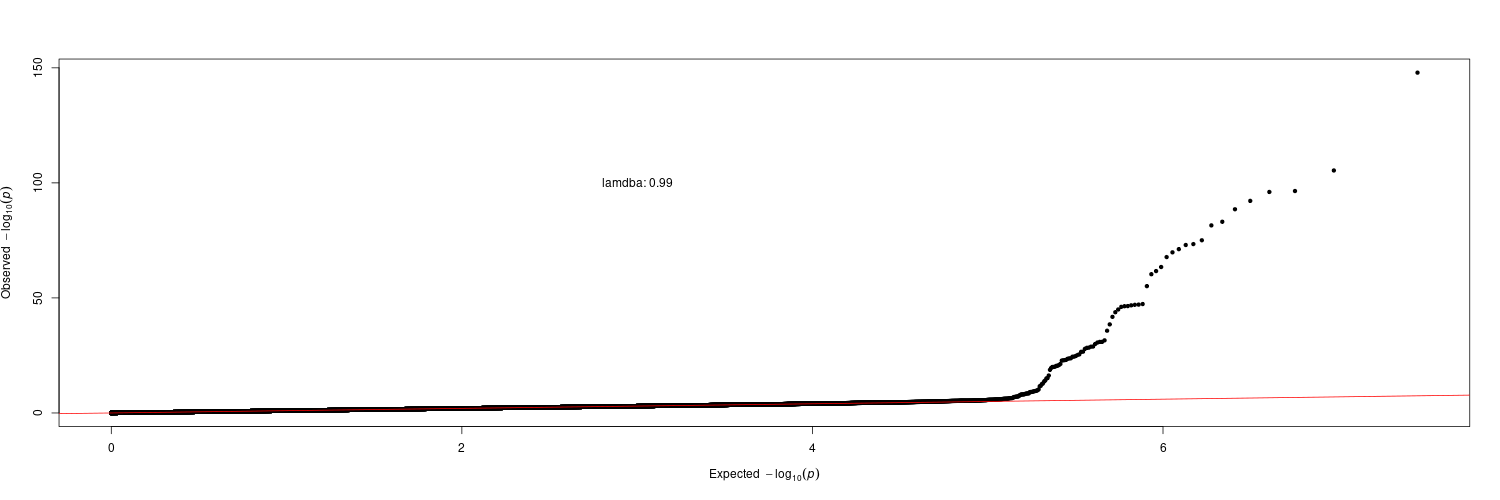

(C) African-American (AA) analysis without APOE E2 and E4 adjustment

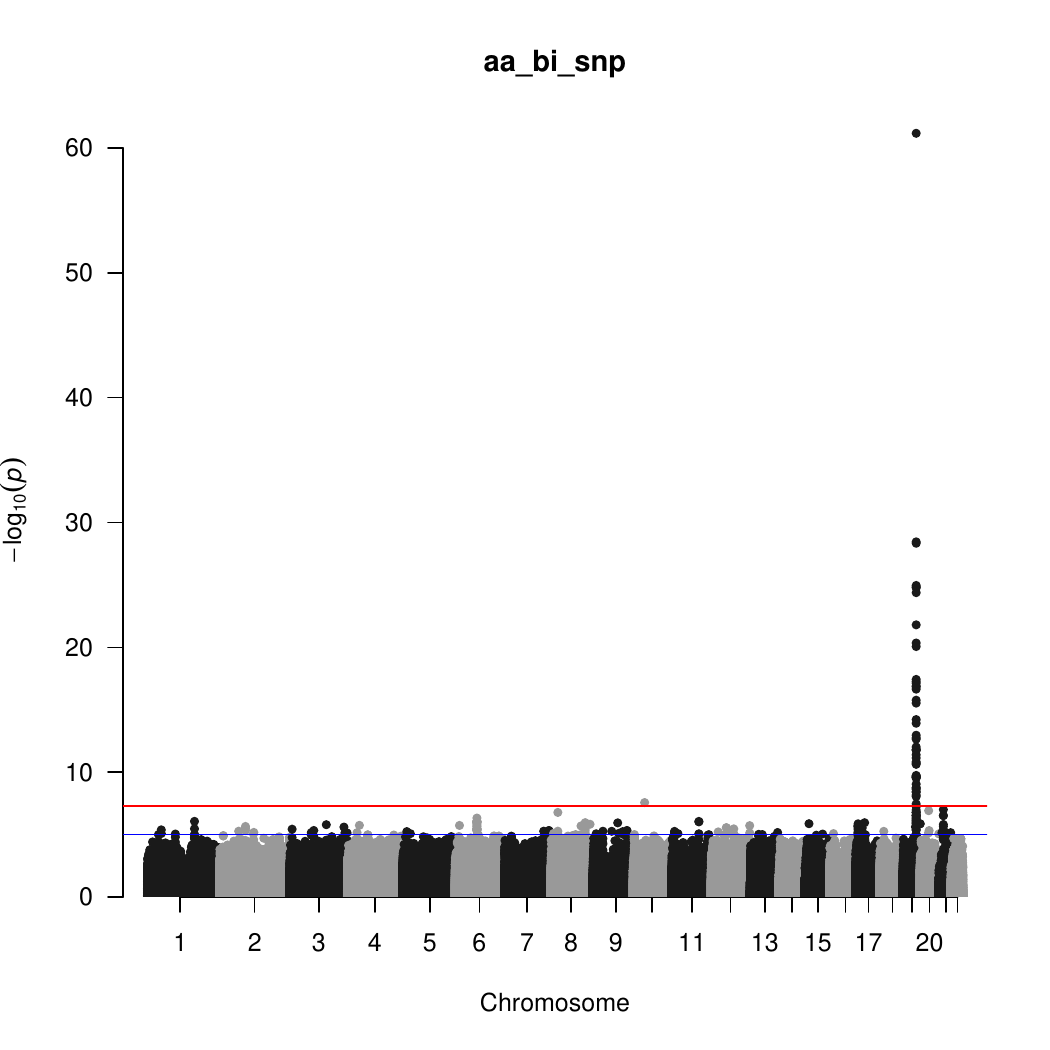

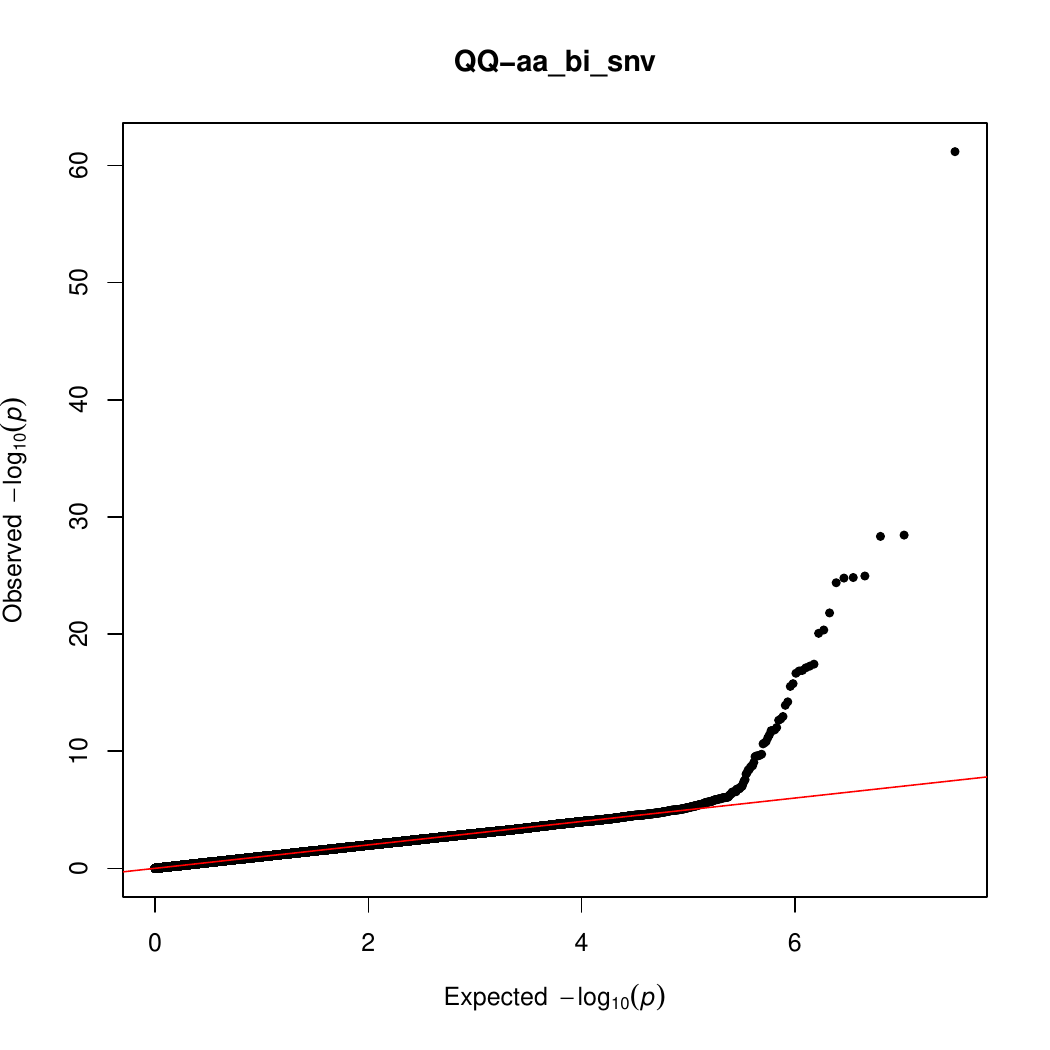

(D) African-American (AA) analysis with APOE E2 and E4 adjustment

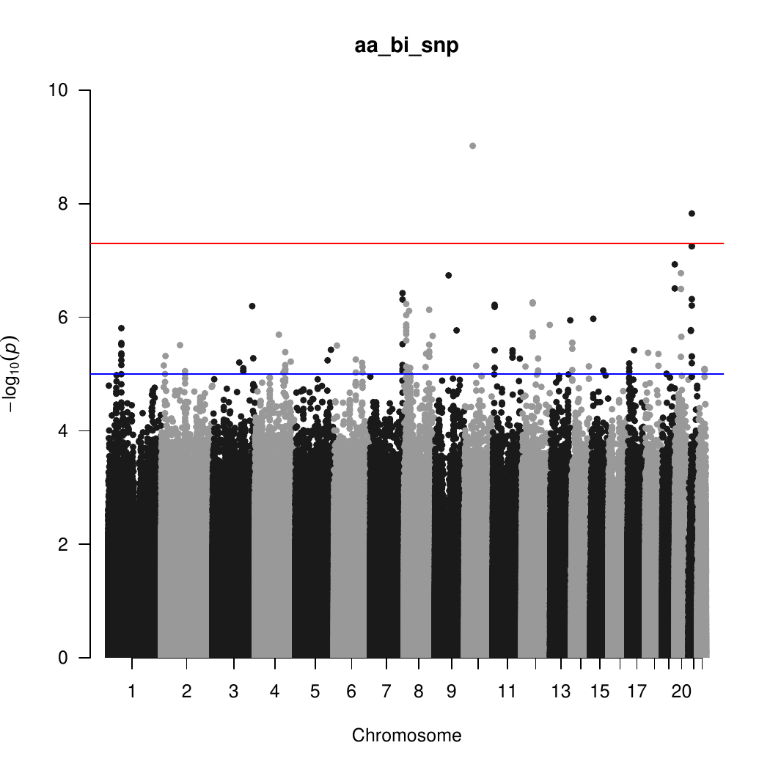

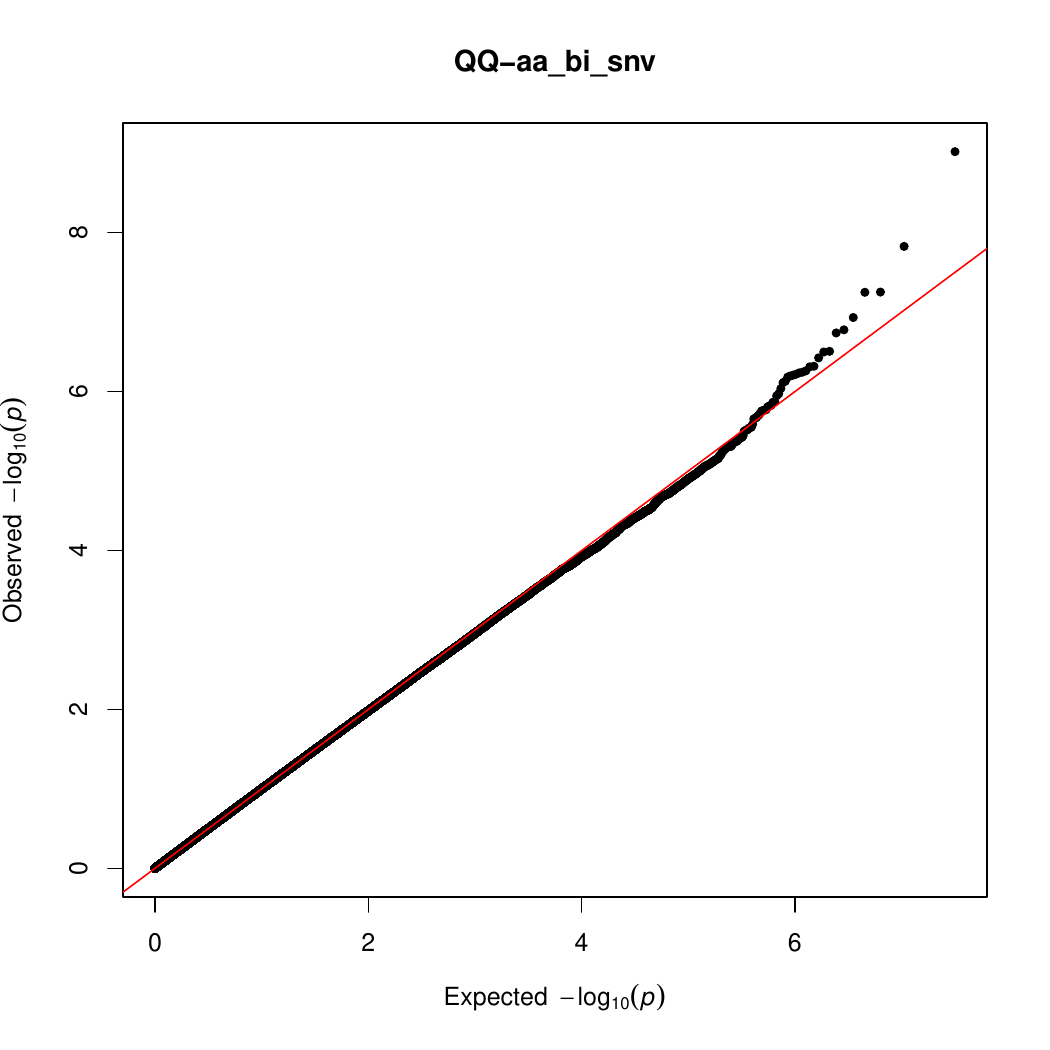

(E) Hispanic (HIS) analysis without APOE E2 and E4 adjustment

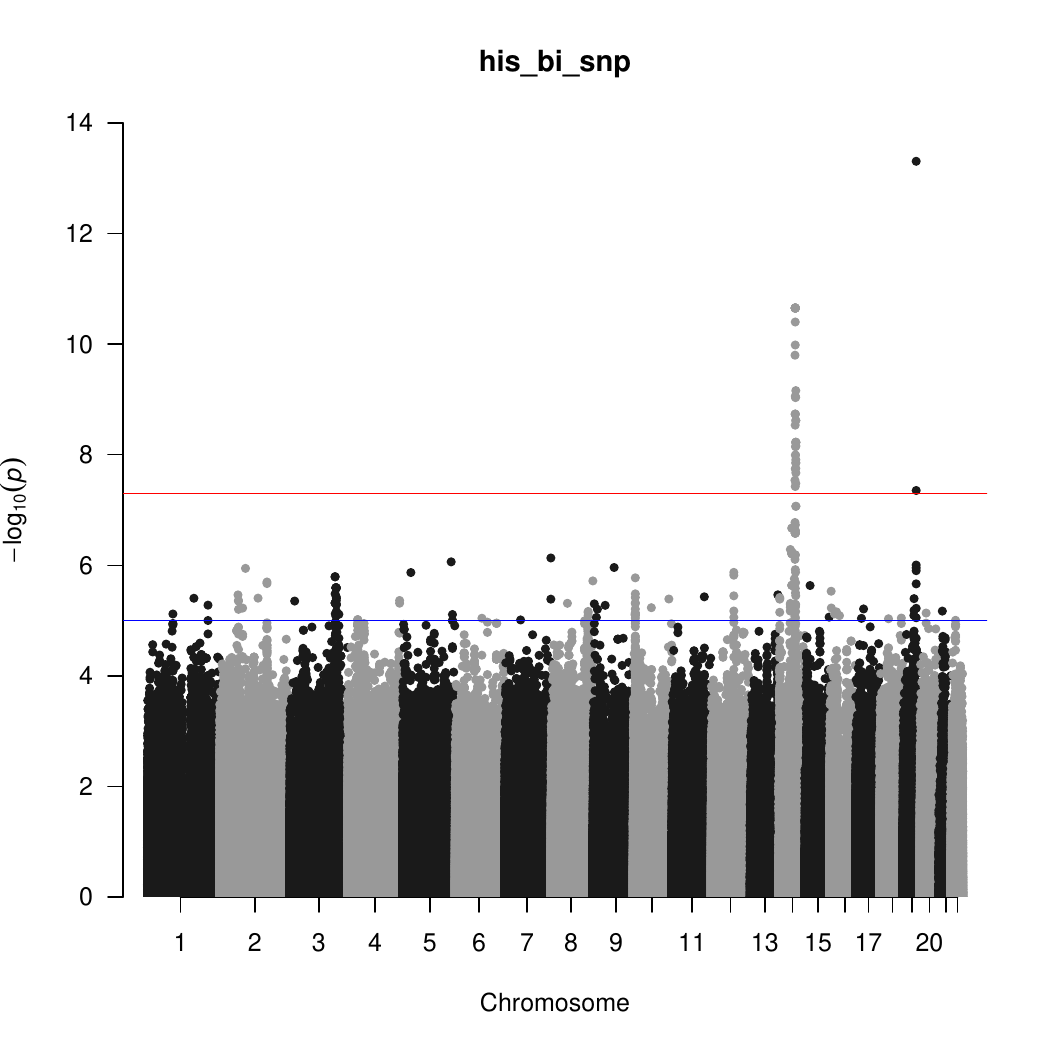

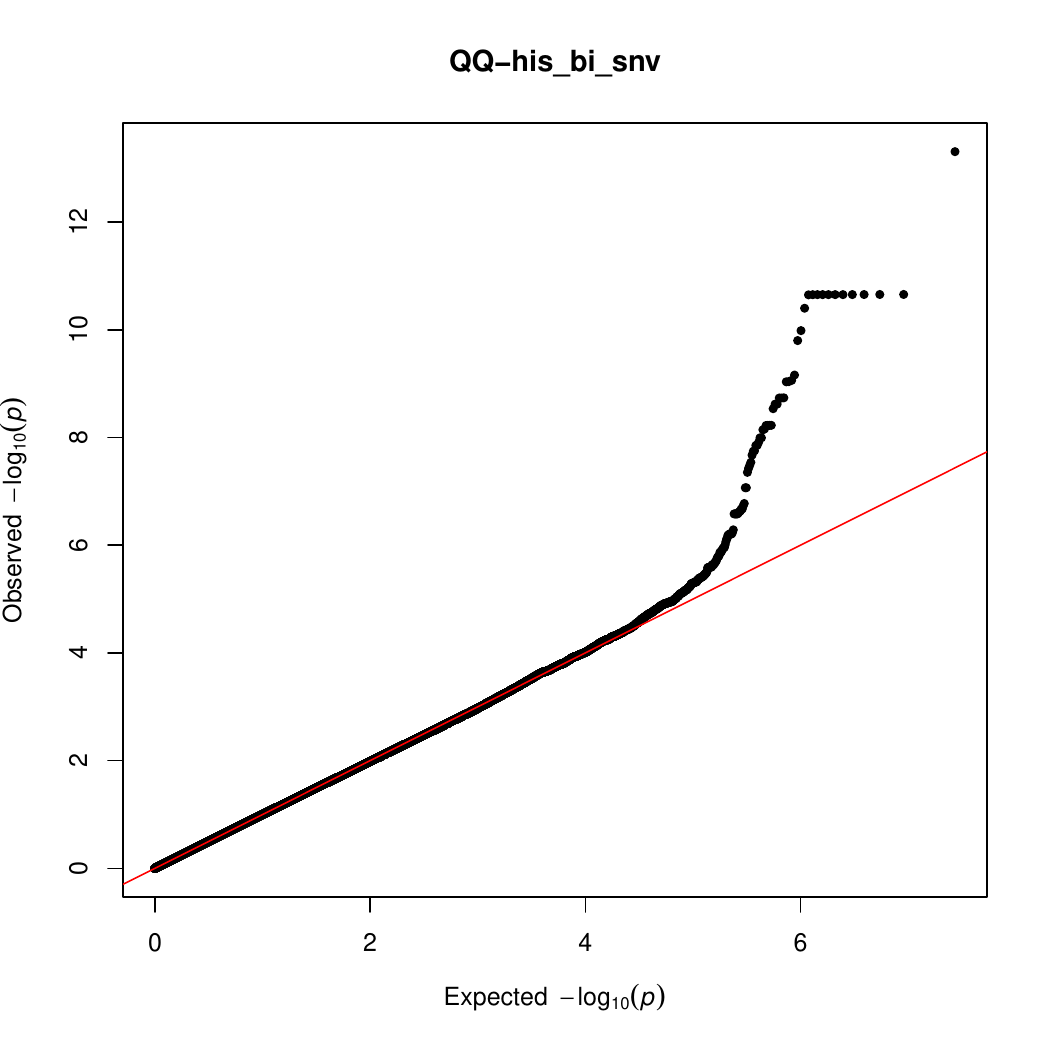

(F) Hispanic (HIS) analysis with APOE E2 and E4 adjustment

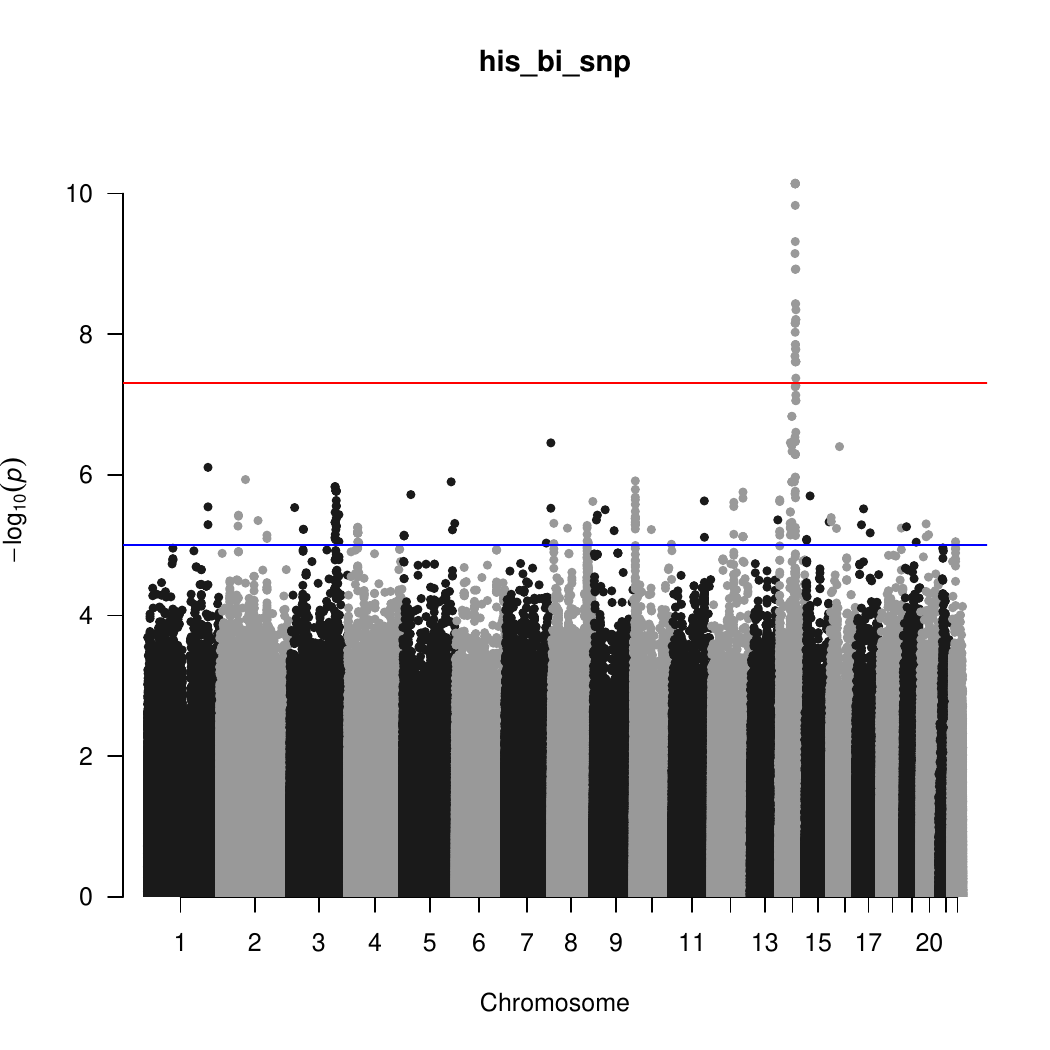

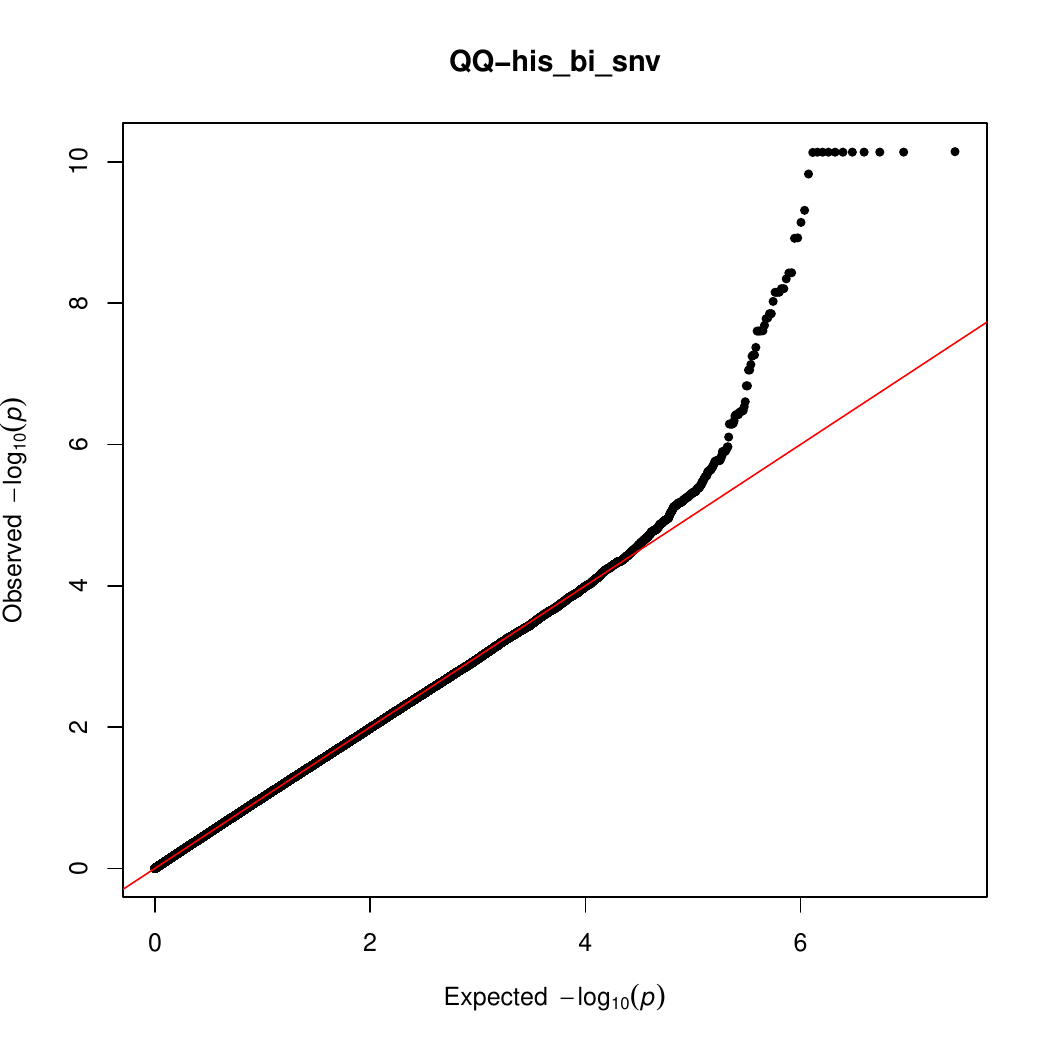

(G) Non-Hispanic White (NHW) analysis without APOE E2 and E4 adjustment

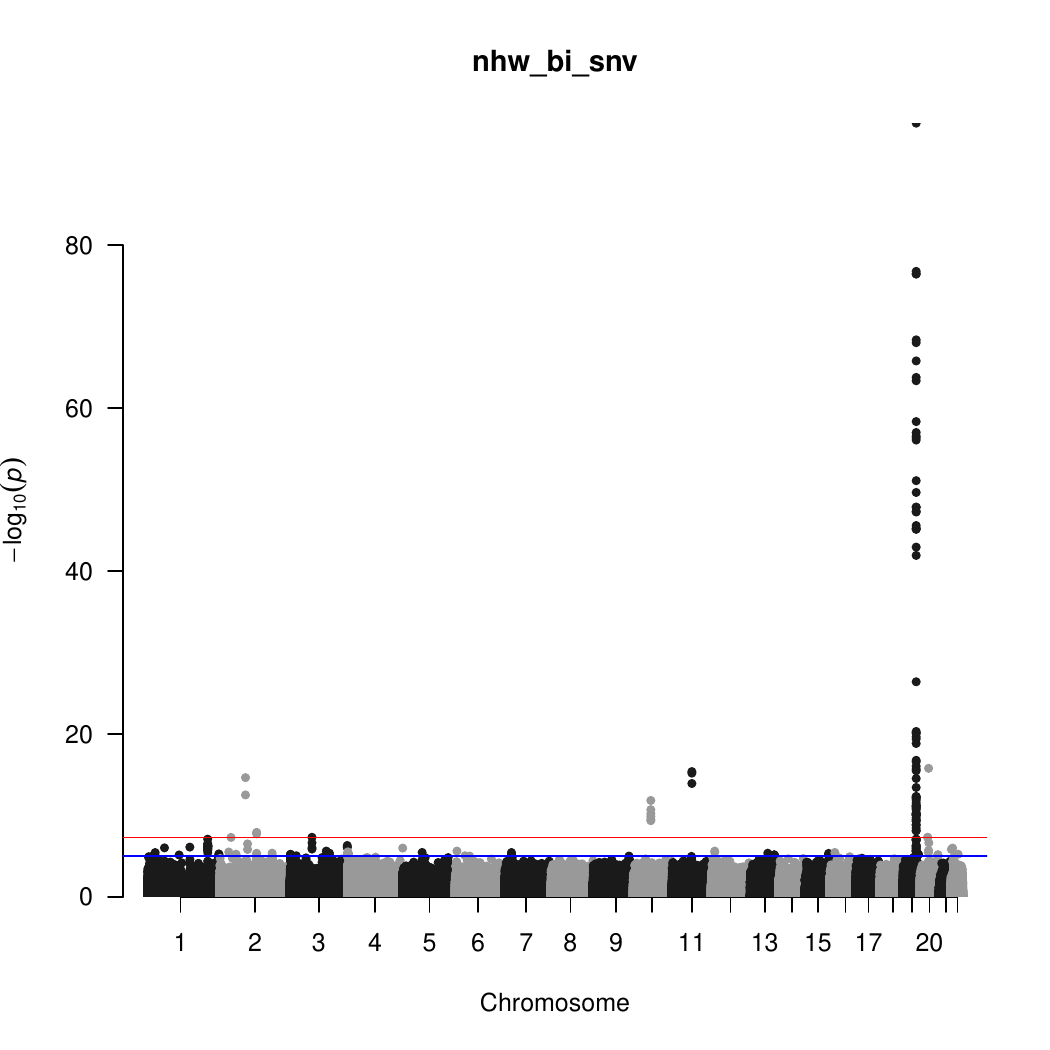

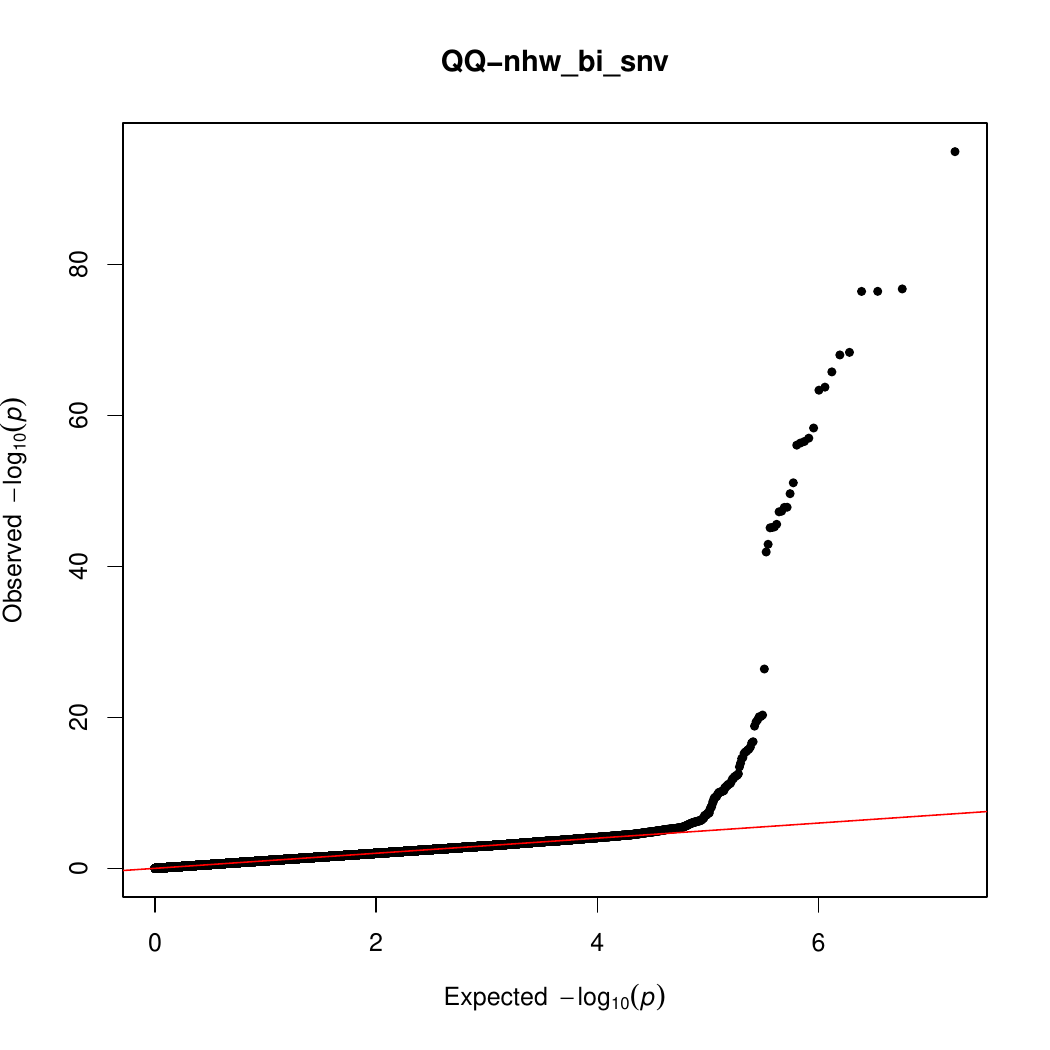

(H) Non-Hispanic White (NHW) analysis with APOE E2 and E4 adjustment

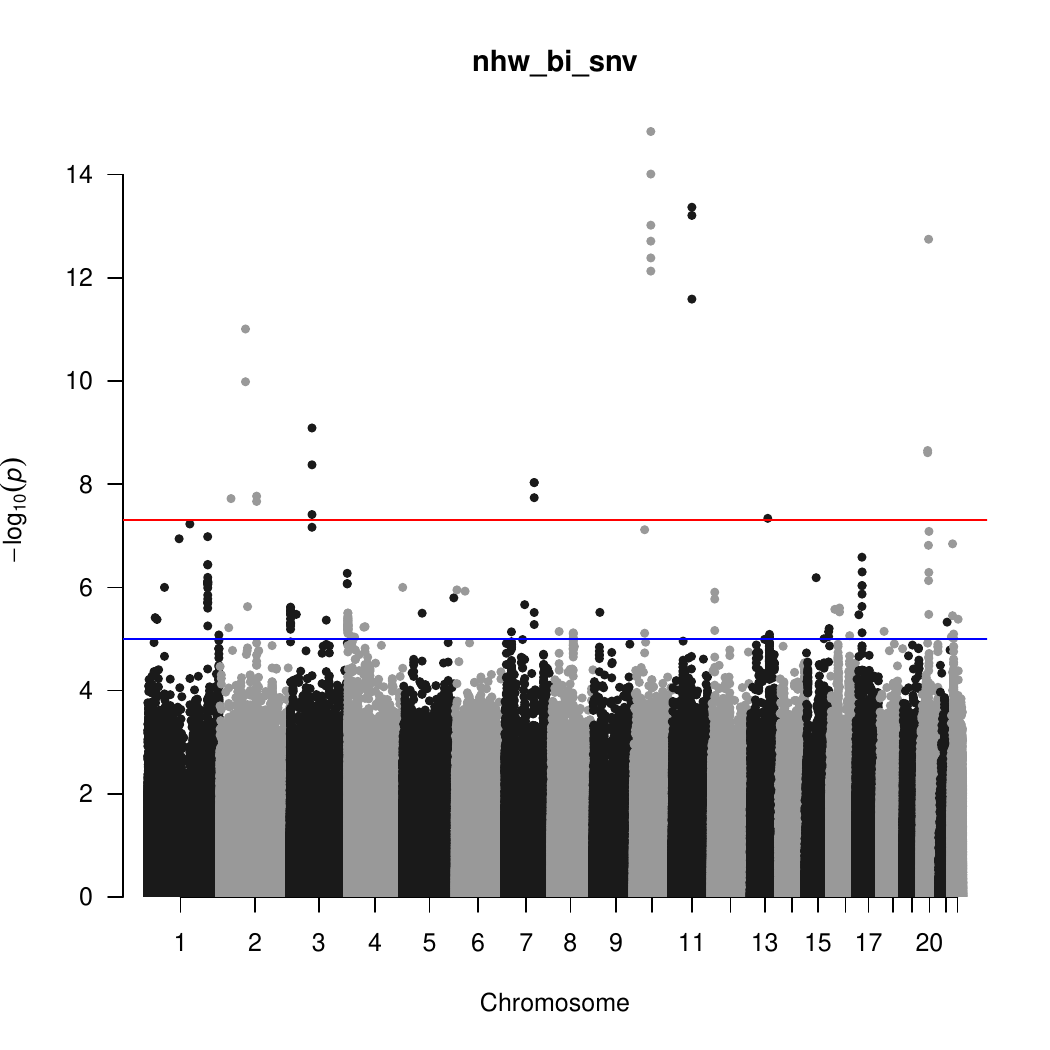

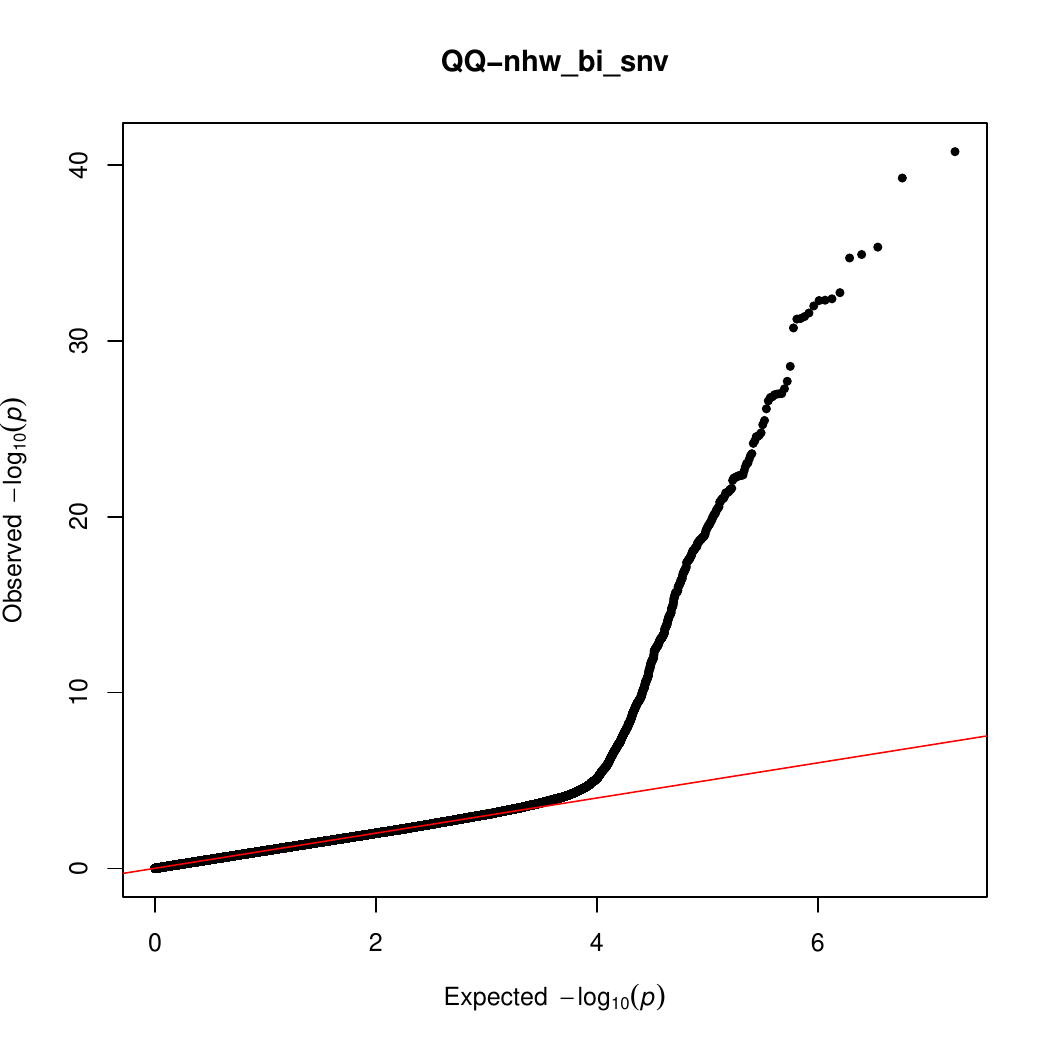

#### Supplementary Figure 2. Regional association plots for genome-wide significant loci.

(A) LINC00320 in AA analysis

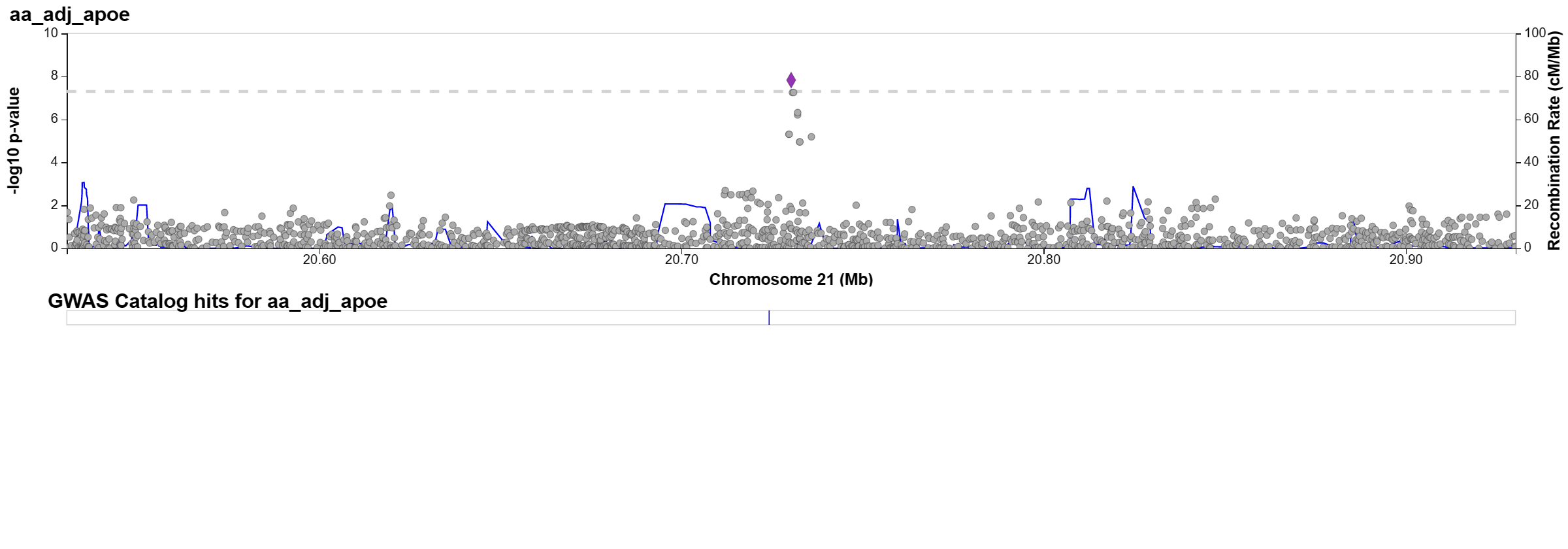

(B) 14q24 in HIS analysis

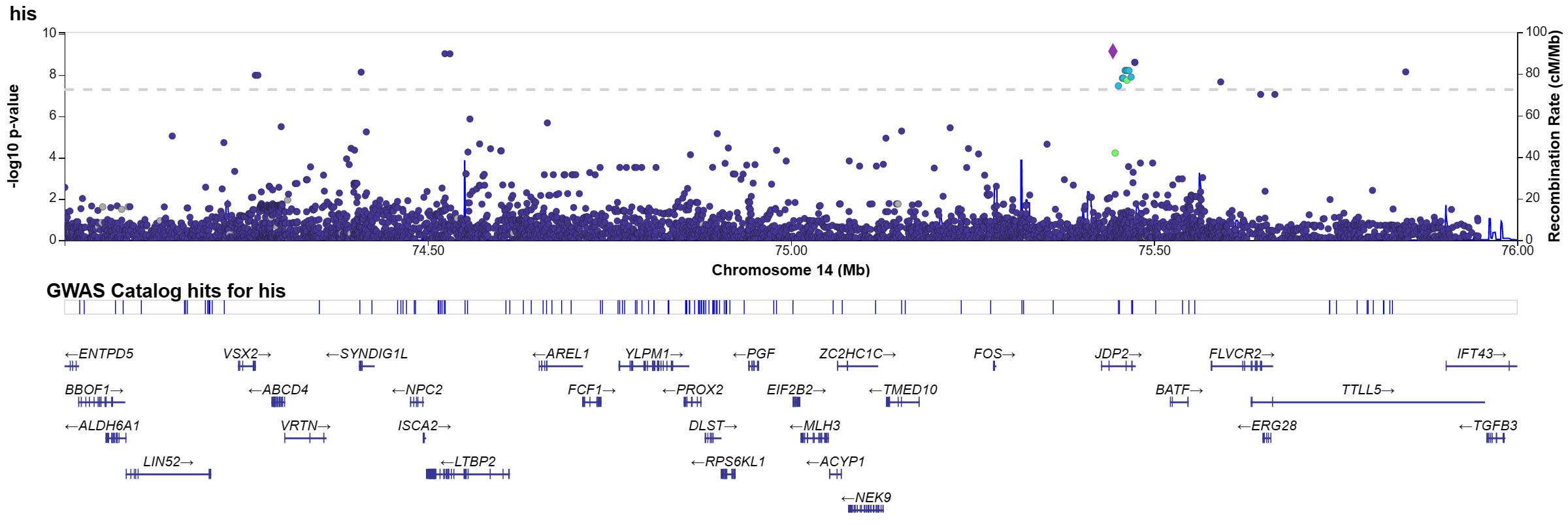

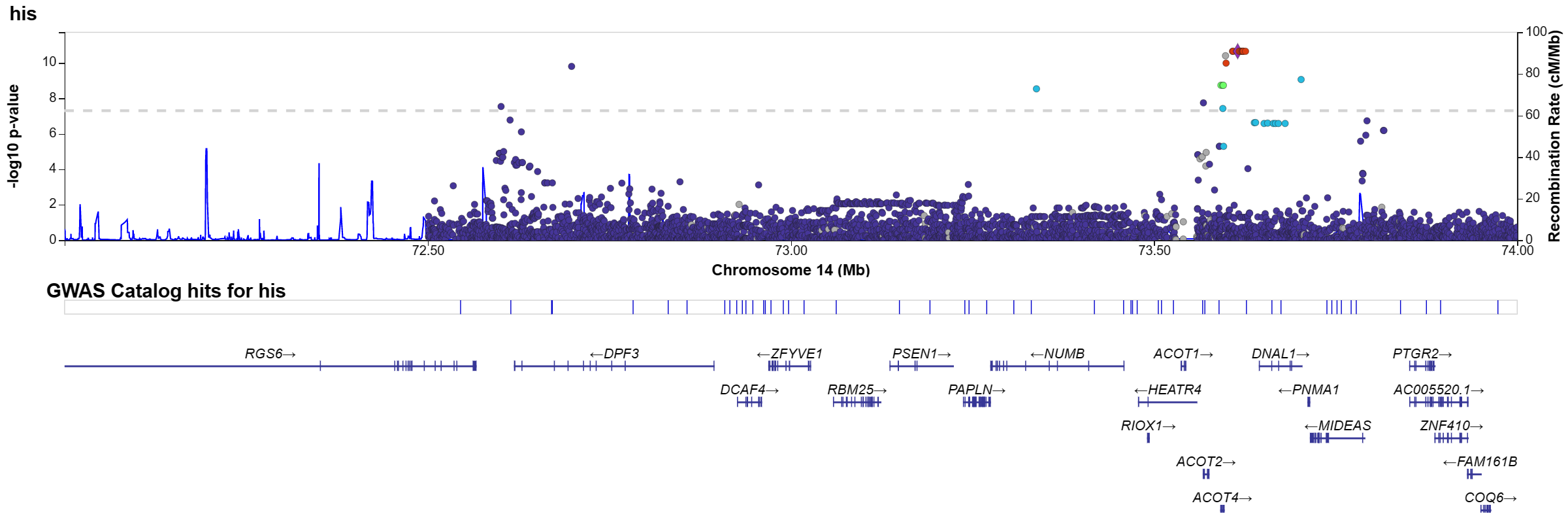

(C) BIN1 in NHW analysis

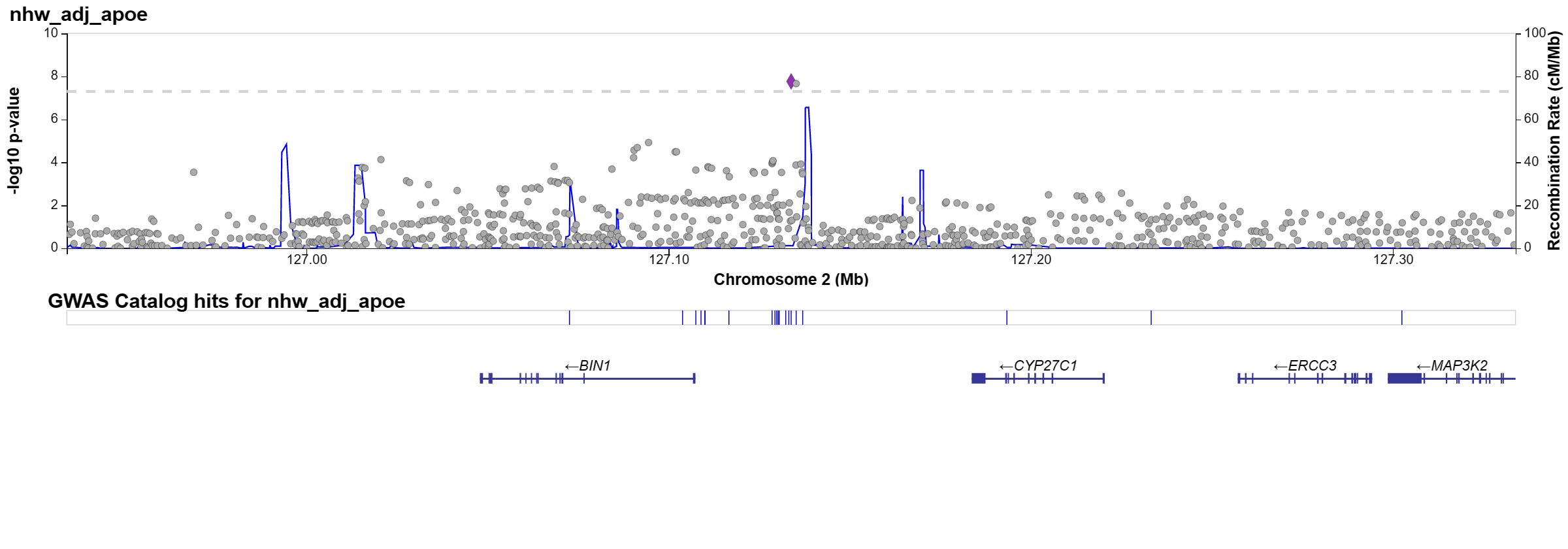

#### Supplementary Figure 3. Kinship coefficient of G206A carriers.

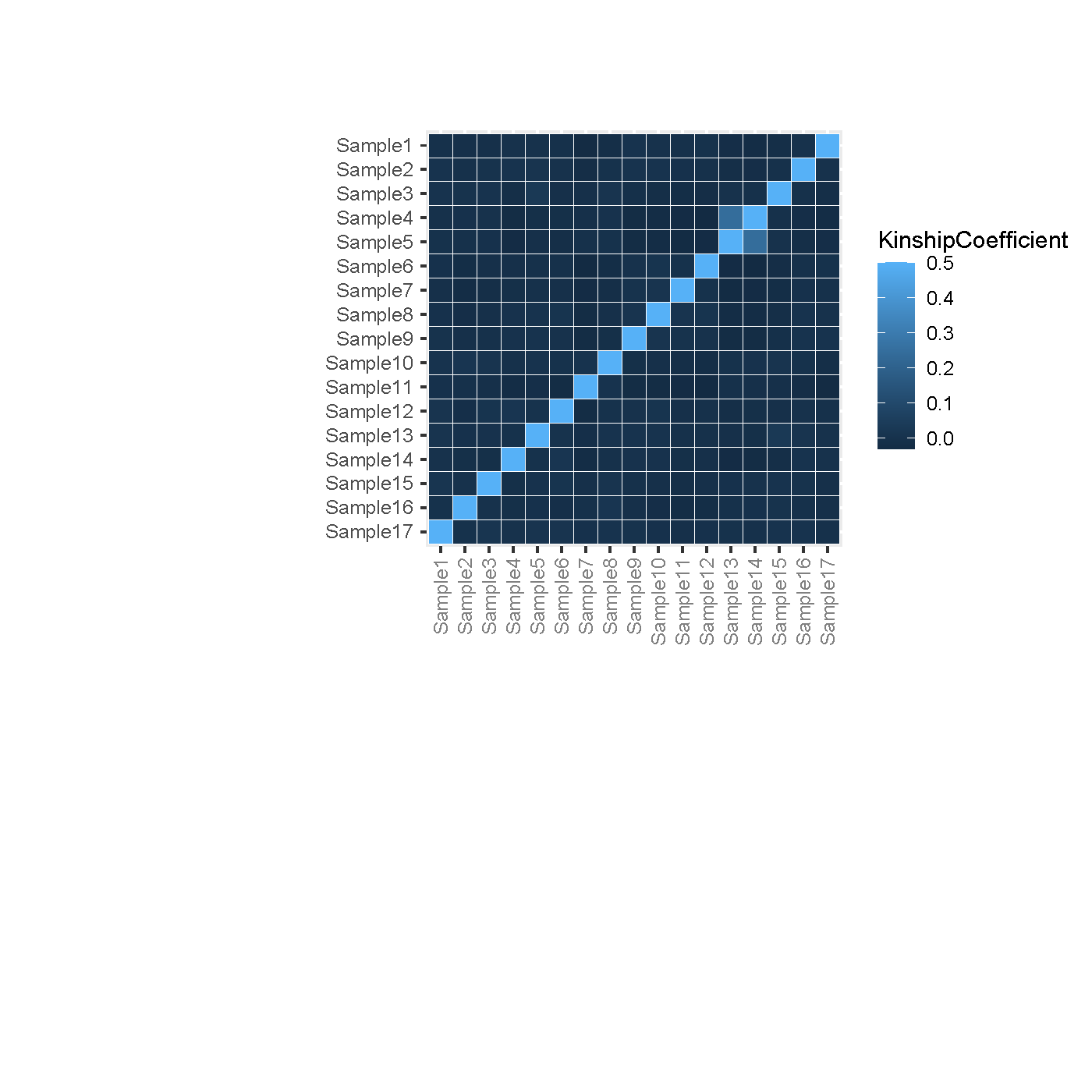

#### Supplementary Figure 4. Principle component analysis of G206A carriers with samples in the 1000 Genomes Project to indicate that G206A carriers are closer to Puerto Rican reference samples (PUR).

The gray-shaded area is the convex hull of the PUR samples, while the black dots indicate the G206A carriers.

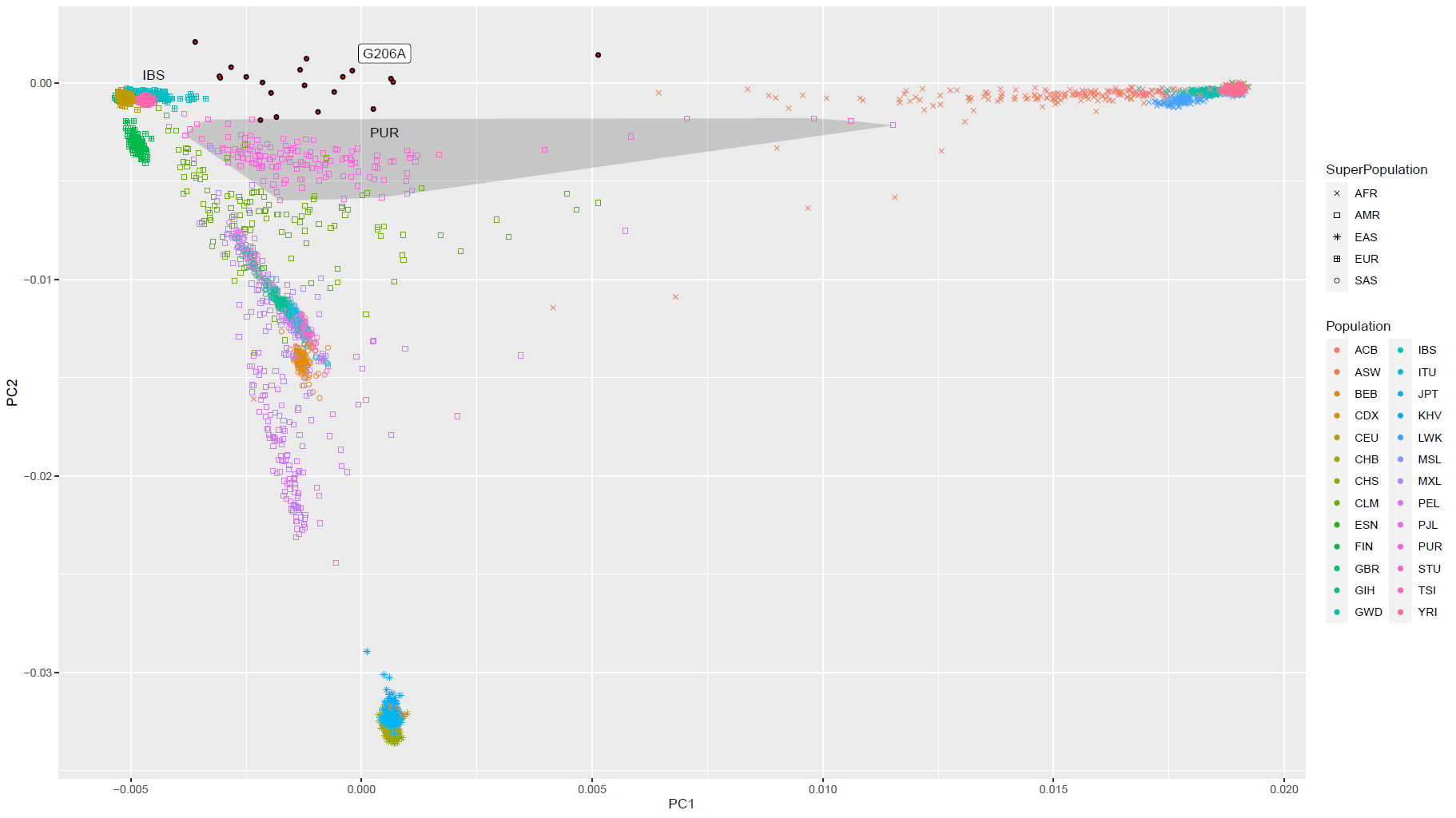

#### Supplementary Figure 5. Principle component analysis of 14q24 mutations carriers with samples in the 1000 Genomes Project to indicate that mutations carriers are closer to Puerto Rican (PUR).

#### Black dots with red * are G206A carriers while black dots with white * are other 14q24 mutation carriers.

The gray-shaded area is the convex hull of PUR samples.

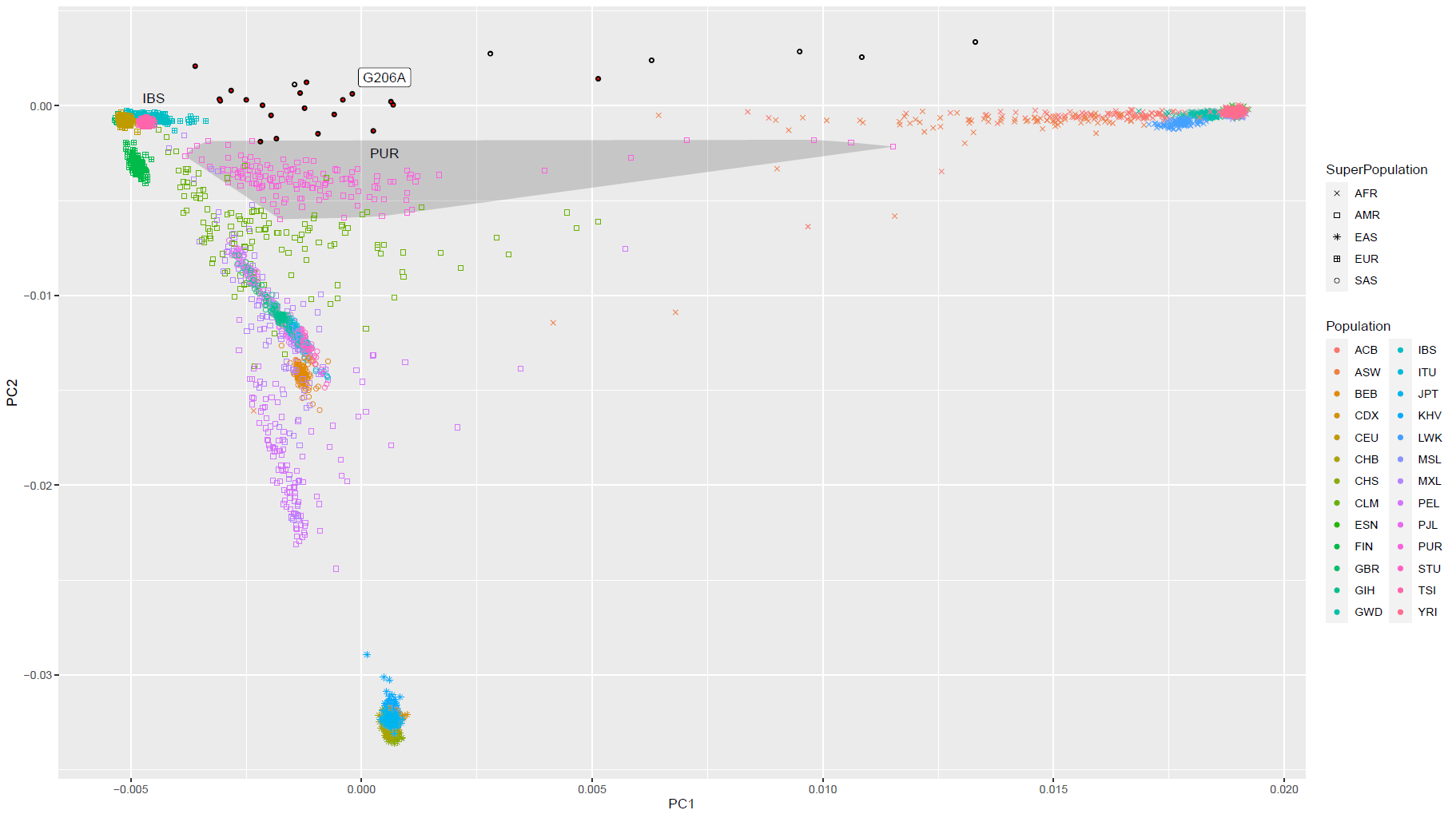

#### Supplementary Figure 6. Local ancestry analysis of G206A carriers to indicate that mutations in 14q24 are based on a founder event of an African.

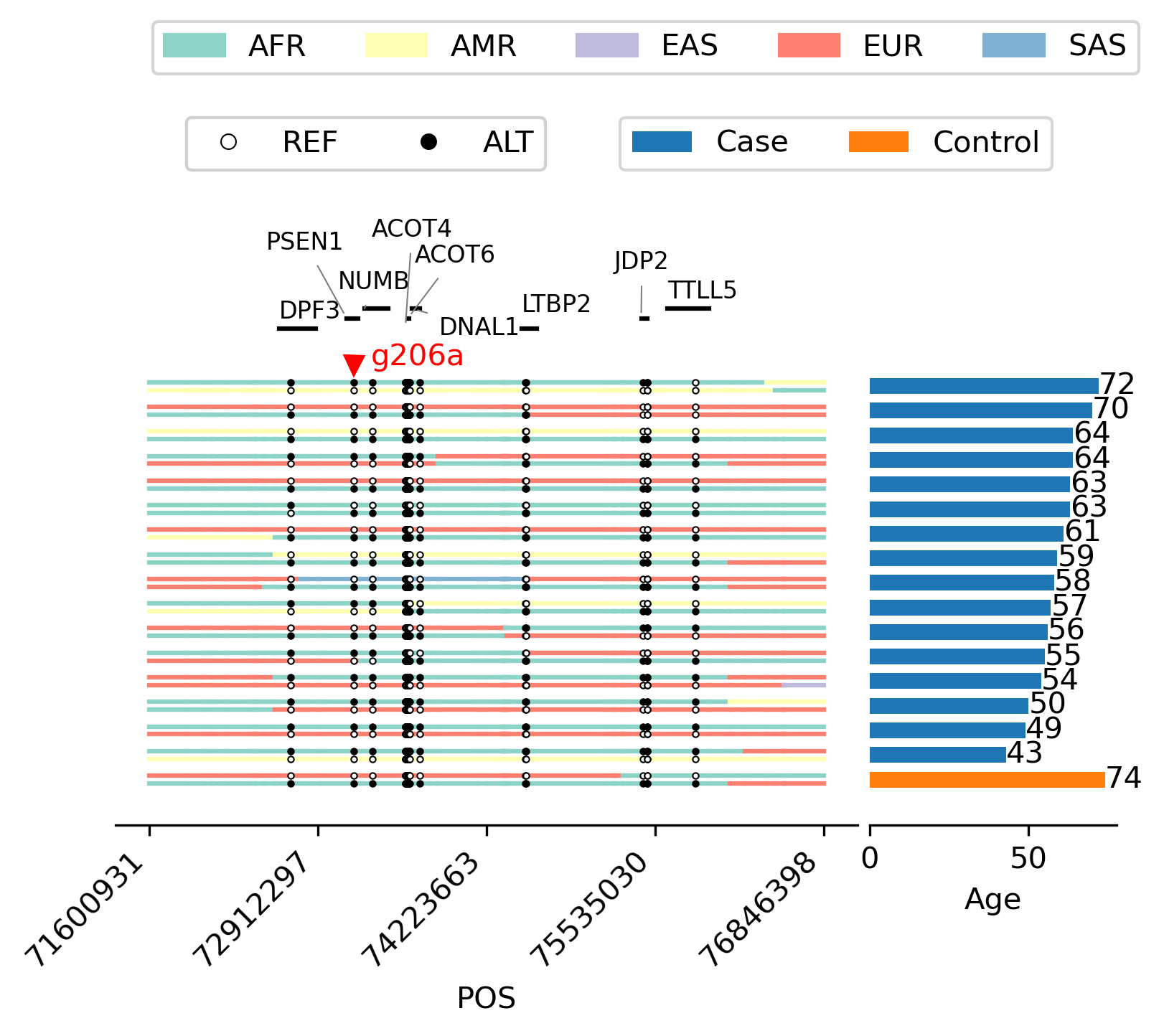

#### Supplementary Figure 7. The IGV views of poorly mapped alignments supporting SNVs in ANK3.

(A) Example Sample1. SNVs are all from supplementary or improper-paired alignments.

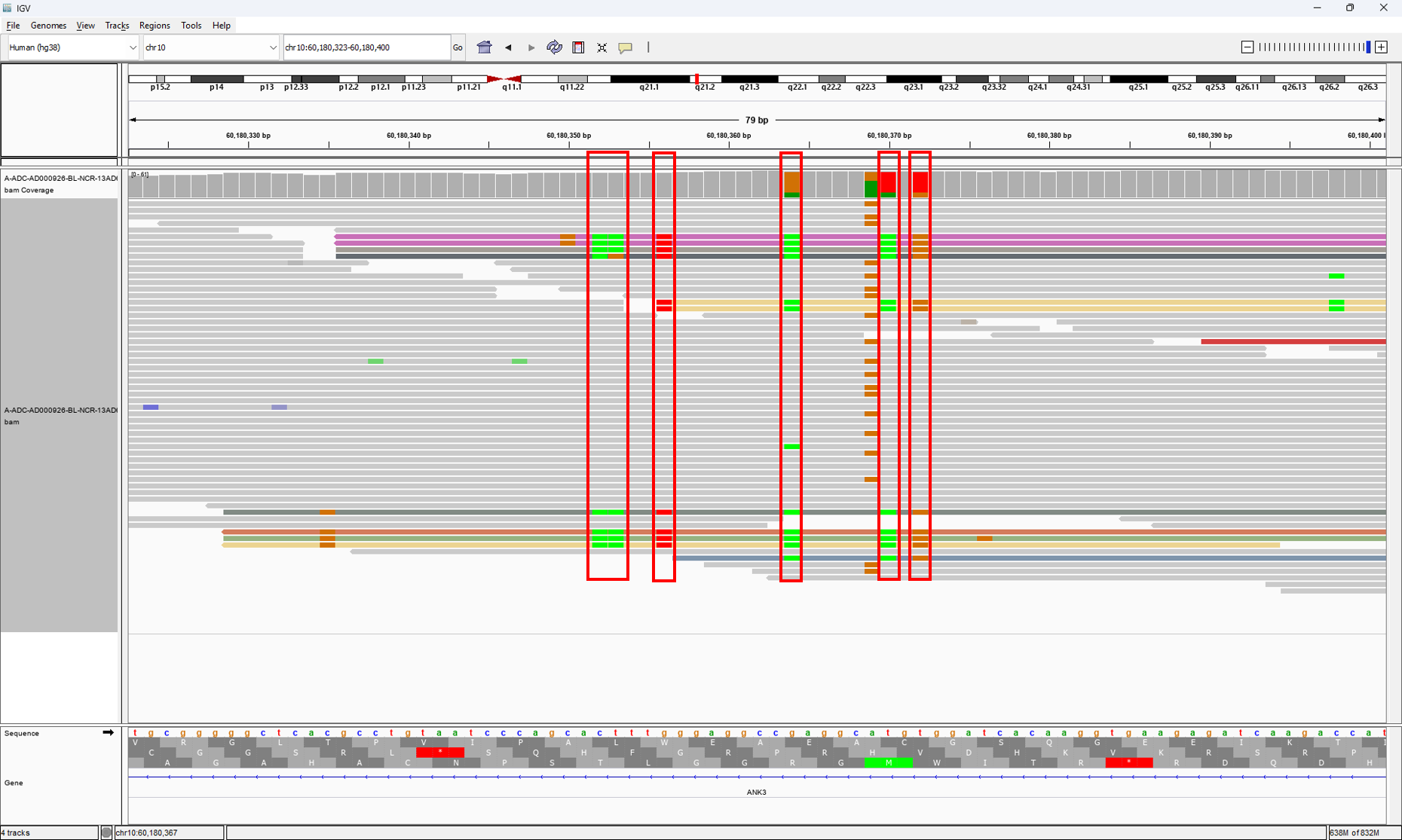

(B) Example Sample2. SNVs are all from supplementary or improper-paired alignments.

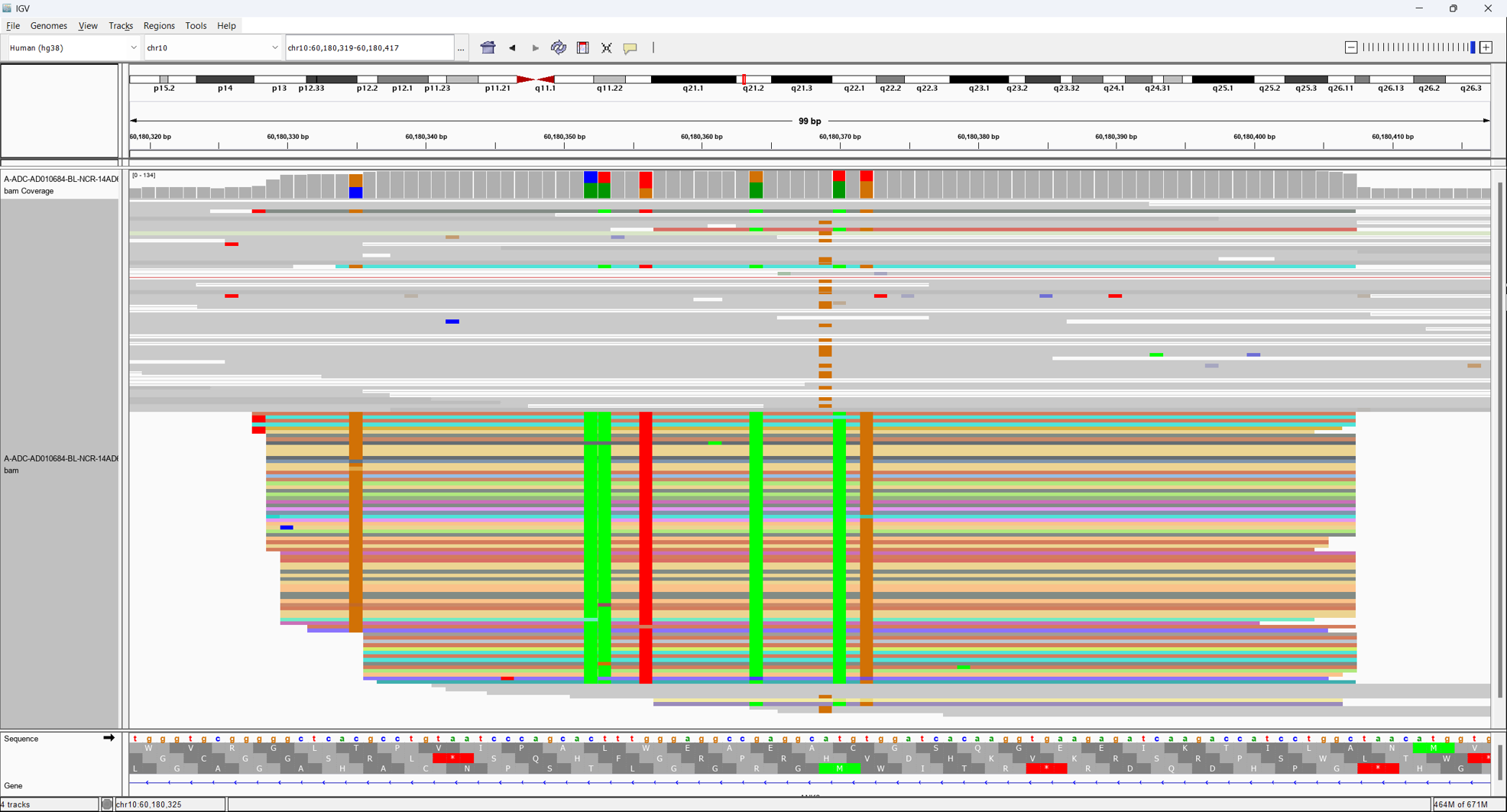

#### Supplementary Figure 8. Euclidean distance of PC1-2 of each our sample with EUR, AFR, and EAS reference populations of the 1000 Genomes Project.

(A) ADSP Non-Hispanic White samples

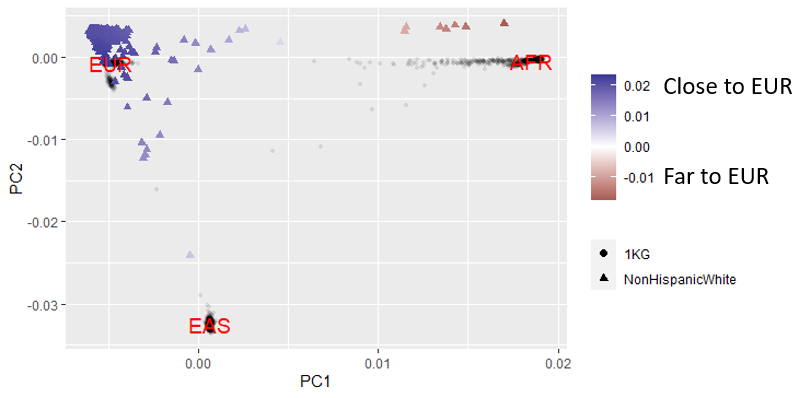

(B) ADSP African American samples

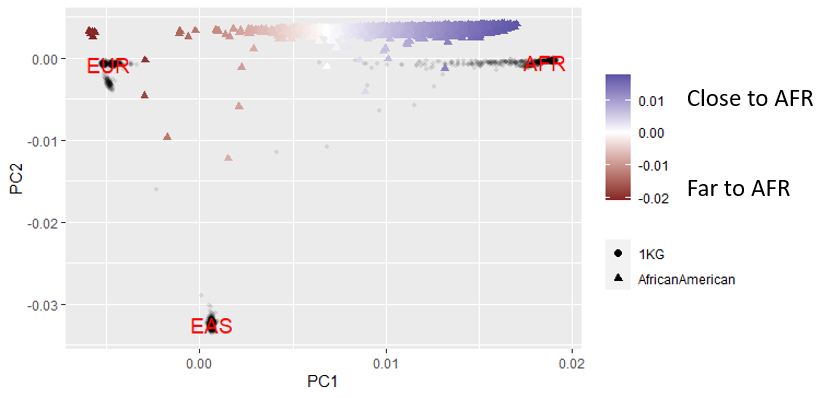

(C) ADSP Hispanic samples

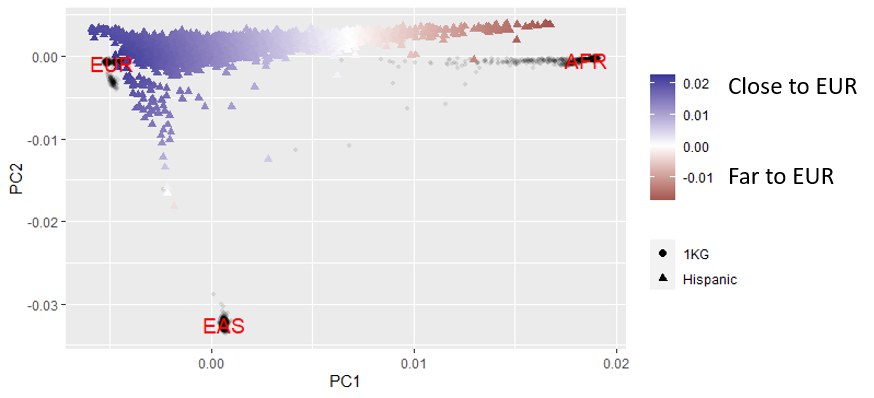

### Supplementary Text

#### Acknowledgements

##### ADSP (sa000001) data:

The Alzheimer’s Disease Sequencing Project (ADSP) is comprised of two Alzheimer’s Disease (AD) genetics consortia and three National Human Genome Research Institute (NHGRI) funded Large Scale Sequencing and Analysis Centers (LSAC). The two AD genetics consortia are the Alzheimer’s Disease Genetics Consortium (ADGC) funded by NIA (U01 AG032984), and the Cohorts for Heart and Aging Research in Genomic Epidemiology (CHARGE) funded by NIA (R01 AG033193), the National Heart, Lung, and Blood Institute (NHLBI), other National Institute of Health (NIH) institutes and other foreign governmental and non-governmental organizations. The Discovery Phase analysis of sequence data is supported through UF1AG047133 (to Drs. Schellenberg, Farrer, Pericak-Vance, Mayeux, and Haines); U01AG049505 to Dr. Seshadri; U01AG049506 to Dr. Boerwinkle; U01AG049507 to Dr. Wijsman; and U01AG049508 to Dr. Goate and the Discovery Extension Phase analysis is supported through U01AG052411 to Dr. Goate, U01AG052410 to Dr. Pericak-Vance and U01 AG052409 to Drs. Seshadri and Fornage.

Sequencing for the Follow Up Study (FUS) is supported through U01AG057659 (to Drs. PericakVance, Mayeux, and Vardarajan) and U01AG062943 (to Drs. Pericak-Vance and Mayeux). Data generation and harmonization in the Follow-up Phase is supported by U54AG052427 (to Drs. Schellenberg and Wang). The FUS Phase analysis of sequence data is supported through U01AG058589 (to Drs. Destefano, Boerwinkle, De Jager, Fornage, Seshadri, and Wijsman), U01AG058654 (to Drs. Haines, Bush, Farrer, Martin, and Pericak-Vance), U01AG058635 (to Dr. Goate), RF1AG058066 (to Drs. Haines, Pericak-Vance, and Scott), RF1AG057519 (to Drs. Farrer and Jun), R01AG048927 (to Dr. Farrer), and RF1AG054074 (to Drs. Pericak-Vance and Beecham).

The ADGC cohorts include: Adult Changes in Thought (ACT) (U01 AG006781, U19 AG066567), the Alzheimer’s Disease Research Centers (ADRC) (P30 AG062429, P30 AG066468, P30 AG062421, P30 AG066509, P30 AG066514, P30 AG066530, P30 AG066507, P30 AG066444, P30 AG066518, P30 AG066512, P30 AG066462, P30 AG072979, P30 AG072972, P30 AG072976, P30 AG072975, P30 AG072978, P30 AG072977, P30 AG066519, P30 AG062677, P30 AG079280, P30 AG062422, P30 AG066511, P30 AG072946, P30 AG062715, P30 AG072973, P30 AG066506, P30 AG066508, P30 AG066515, P30 AG072947, P30 AG072931, P30 AG066546, P20 AG068024, P20 AG068053, P20 AG068077, P20 AG068082, P30 AG072958, P30 AG072959), the Chicago Health and Aging Project (CHAP) (R01 AG11101, RC4 AG039085, K23 AG030944), Indiana Memory and Aging Study (IMAS) (R01 AG019771), Indianapolis Ibadan (R01 AG009956, P30 AG010133), the Memory and Aging Project (MAP) ( R01 AG17917), Mayo Clinic (MAYO) (R01 AG032990, U01 AG046139, R01 NS080820, RF1 AG051504, P50 AG016574), Mayo Parkinson’s Disease controls (NS039764, NS071674, 5RC2HG005605), University of Miami (R01 AG027944, R01 AG028786, R01 AG019085, IIRG09133827, A2011048), the Multi-Institutional Research in Alzheimer’s Genetic Epidemiology Study (MIRAGE) (R01 AG09029, R01 AG025259), the National Centralized Repository for Alzheimer’s Disease and Related Dementias (NCRAD) (U24 AG021886), the National Institute on Aging Late Onset Alzheimer’s Disease Family Study (NIA- LOAD) (U24 AG056270), the Religious Orders Study (ROS) (P30 AG10161, R01 AG15819), the Texas Alzheimer’s Research and Care Consortium (TARCC) (funded by the Darrell K Royal Texas Alzheimer’s Initiative), Vanderbilt University/Case Western Reserve University (VAN/CWRU) (R01 AG019757, R01 AG021547, R01 AG027944, R01 AG028786, P01 NS026630, and Alzheimer’s Association), the Washington Heights-Inwood Columbia Aging Project (WHICAP) (RF1 AG054023), the University of Washington Families (VA Research Merit Grant, NIA: P50AG005136, R01AG041797, NINDS: R01NS069719), the Columbia University Hispanic Estudio Familiar de Influencia Genetica de Alzheimer (EFIGA) (RF1 AG015473), the University of Toronto (UT) (funded by Wellcome Trust, Medical Research Council, Canadian Institutes of Health Research), and Genetic Differences (GD) (R01 AG007584). The CHARGE cohorts are supported in part by National Heart, Lung, and Blood Institute (NHLBI) infrastructure grant HL105756 (Psaty), RC2HL102419 (Boerwinkle) and the neurology working group is supported by the National Institute on Aging (NIA) R01 grant AG033193.

The CHARGE cohorts participating in the ADSP include the following: Austrian Stroke Prevention Study (ASPS), ASPS-Family study, and the Prospective Dementia Registry-Austria (ASPS/PRODEM-Aus), the Atherosclerosis Risk in Communities (ARIC) Study, the Cardiovascular Health Study (CHS), the Erasmus Rucphen Family Study (ERF), the Framingham Heart Study (FHS), and the Rotterdam Study (RS). ASPS is funded by the Austrian Science Fond (FWF) grant number P20545-P05 and P13180 and the Medical University of Graz. The ASPS-Fam is funded by the Austrian Science Fund (FWF) project I904), the EU Joint Programme – Neurodegenerative Disease Research (JPND) in frame of the BRIDGET project (Austria, Ministry of Science) and the Medical University of Graz and the Steiermärkische Krankenanstalten Gesellschaft. PRODEM-Austria is supported by the Austrian Research Promotion agency (FFG) (Project No. 827462) and by the Austrian National Bank (Anniversary Fund, project 15435. ARIC research is carried out as a collaborative study supported by NHLBI contracts (HHSN268201100005C, HHSN268201100006C, HHSN268201100007C, HHSN268201100008C, HHSN268201100009C, HHSN268201100010C, HHSN268201100011C, and HHSN268201100012C). Neurocognitive data in ARIC is collected by U01 2U01HL096812, 2U01HL096814, 2U01HL096899, 2U01HL096902, 2U01HL096917 from the NIH (NHLBI, NINDS, NIA and NIDCD), and with previous brain MRI examinations funded by R01-HL70825 from the NHLBI. CHS research was supported by contracts HHSN268201200036C, HHSN268200800007C, N01HC55222, N01HC85079, N01HC85080, N01HC85081, N01HC85082, N01HC85083, N01HC85086, and grants U01HL080295 and U01HL130114 from the NHLBI with additional contribution from the National Institute of Neurological Disorders and Stroke (NINDS). Additional support was provided by R01AG023629, R01AG15928, and R01AG20098 from the NIA. FHS research is supported by NHLBI contracts N01-HC-25195 and HHSN268201500001I. This study was also supported by additional grants from the NIA (R01s AG054076, AG049607 and AG033040 and NINDS (R01 NS017950). The ERF study as a part of EUROSPAN (European Special Populations Research Network) was supported by European Commission FP6 STRP grant number 018947 (LSHG-CT-2006-01947) and also received funding from the European Community’s Seventh Framework Programme (FP7/2007-2013)/grant agreement HEALTH-F4- 2007-201413 by the European Commission under the programme “Quality of Life and Management of the Living Resources” of 5th Framework Programme (no. QLG2-CT-2002- 01254). High-throughput analysis of the ERF data was supported by a joint grant from the Netherlands Organization for Scientific Research and the Russian Foundation for Basic Research (NWO-RFBR 047.017.043). The Rotterdam Study is funded by Erasmus Medical Center and Erasmus University, Rotterdam, the Netherlands Organization for Health Research and Development (ZonMw), the Research Institute for Diseases in the Elderly (RIDE), the Ministry of Education, Culture and Science, the Ministry for Health, Welfare and Sports, the European Commission (DG XII), and the municipality of Rotterdam. Genetic data sets are also supported by the Netherlands Organization of Scientific Research NWO Investments (175.010.2005.011, 911-03-012), the Genetic Laboratory of the Department of Internal Medicine, Erasmus MC, the Research Institute for Diseases in the Elderly (014-93-015; RIDE2), and the Netherlands Genomics Initiative (NGI)/Netherlands Organization for Scientific Research (NWO) Netherlands Consortium for Healthy Aging (NCHA), project 050-060-810. All studies are grateful to their participants, faculty and staff. The content of these manuscripts is solely the responsibility of the authors and does not necessarily represent the official views of the National Institutes of Health or the U.S. Department of Health and Human Services.

The FUS cohorts include: the Alzheimer’s Disease Research Centers (ADRC) (P30 AG062429, P30 AG066468, P30 AG062421, P30 AG066509, P30 AG066514, P30 AG066530, P30 AG066507, P30 AG066444, P30 AG066518, P30 AG066512, P30 AG066462, P30 AG072979, P30 AG072972, P30 AG072976, P30 AG072975, P30 AG072978, P30 AG072977, P30 AG066519, P30 AG062677, P30 AG079280, P30 AG062422, P30 AG066511, P30 AG072946, P30 AG062715, P30 AG072973, P30 AG066506, P30 AG066508, P30 AG066515, P30 AG072947, P30 AG072931, P30 AG066546, P20 AG068024, P20 AG068053, P20 AG068077, P20 AG068082, P30 AG072958, P30 AG072959), Alzheimer’s Disease Neuroimaging Initiative (ADNI) (U19AG024904), Amish Protective Variant Study (RF1AG058066), Cache County Study (R01AG11380, R01AG031272, R01AG21136, RF1AG054052), Case Western Reserve University Brain Bank (CWRUBB) (P50AG008012), Case Western Reserve University Rapid Decline (CWRURD) (RF1AG058267, NU38CK000480), CubanAmerican Alzheimer’s Disease Initiative (CuAADI) (3U01AG052410), Estudio Familiar de Influencia Genetica en Alzheimer (EFIGA) (5R37AG015473, RF1AG015473, R56AG051876), Genetic and Environmental Risk Factors for Alzheimer Disease Among African Americans Study (GenerAAtions) (2R01AG09029, R01AG025259, 2R01AG048927), Gwangju Alzheimer and Related Dementias Study (GARD) (U01AG062602), Hillblom Aging Network (2014-A-004-NET, R01AG032289, R01AG048234), Hussman Institute for Human Genomics Brain Bank (HIHGBB) (R01AG027944, Alzheimer’s Association “Identification of Rare Variants in Alzheimer Disease”), Ibadan Study of Aging (IBADAN) (5R01AG009956), Longevity Genes Project (LGP) and LonGenity (R01AG042188, R01AG044829, R01AG046949, R01AG057909, R01AG061155, P30AG038072), Mexican Health and Aging Study (MHAS) (R01AG018016), Multi-Institutional Research in Alzheimer’s Genetic Epidemiology (MIRAGE) (2R01AG09029, R01AG025259, 2R01AG048927), Northern Manhattan Study (NOMAS) (R01NS29993), Peru Alzheimer’s Disease Initiative (PeADI) (RF1AG054074), Puerto Rican 1066 (PR1066) (Wellcome Trust (GR066133/GR080002), European Research Council (340755)), Puerto Rican Alzheimer Disease Initiative (PRADI) (RF1AG054074), Reasons for Geographic and Racial Differences in Stroke (REGARDS) (U01NS041588), Research in African American Alzheimer Disease Initiative (REAAADI) (U01AG052410), the Religious Orders Study (ROS) (P30 AG10161, P30 AG72975, R01 AG15819, R01 AG42210), the RUSH Memory and Aging Project (MAP) (R01 AG017917, R01 AG42210Stanford Extreme Phenotypes in AD (R01AG060747), University of Miami Brain Endowment Bank (MBB), University of Miami/Case Western/North Carolina A&T African American (UM/CASE/NCAT) (U01AG052410, R01AG028786), and Wisconsin Registry for Alzheimer’s Prevention (WRAP) (R01AG027161 and R01AG054047).

The four LSACs are: the Human Genome Sequencing Center at the Baylor College of Medicine (U54 HG003273), the Broad Institute Genome Center (U54HG003067), The American Genome Center at the Uniformed Services University of the Health Sciences (U01AG057659), and the Washington University Genome Institute (U54HG003079). Genotyping and sequencing for the ADSP FUS is also conducted at John P. Hussman Institute for Human Genomics (HIHG) Center for Genome Technology (CGT).

Biological samples and associated phenotypic data used in primary data analyses were stored at Study Investigators institutions, and at the National Centralized Repository for Alzheimer’s Disease and Related Dementias (NCRAD, U24AG021886) at Indiana University funded by NIA. Associated Phenotypic Data used in primary and secondary data analyses were provided by Study Investigators, the NIA funded Alzheimer’s Disease Centers (ADCs), and the National Alzheimer’s Coordinating Center (NACC, U24AG072122) and the National Institute on Aging Genetics of Alzheimer’s Disease Data Storage Site (NIAGADS, U24AG041689) at the University of Pennsylvania, funded by NIA. Harmonized phenotypes were provided by the ADSP Phenotype Harmonization Consortium (ADSP-PHC), funded by NIA (U24 AG074855, U01 AG068057 and R01 AG059716) and Ultrascale Machine Learning to Empower Discovery in Alzheimer’s Disease Biobanks (AI4AD, U01 AG068057). This research was supported in part by the Intramural Research Program of the National Institutes of health, National Library of Medicine. Contributors to the Genetic Analysis Data included Study Investigators on projects that were individually funded by NIA, and other NIH institutes, and by private U.S. organizations, or foreign governmental or nongovernmental organizations.

##### ADNI (sa000002) data:

Data collection and sharing for this project was funded by the Alzheimer's Disease Neuroimaging Initiative (ADNI) (National Institutes of Health Grant U01 AG024904) and DOD ADNI (Department of Defense award number W81XWH-12-2-0012). ADNI is funded by the National Institute on Aging, the National Institute of Biomedical Imaging and Bioengineering, and through generous contributions from the following: AbbVie, Alzheimer’s Association; Alzheimer’s Drug Discovery Foundation; Araclon Biotech; BioClinica, Inc.; Biogen; Bristol-Myers Squibb Company; CereSpir, Inc.; Cogstate; Eisai Inc.; Elan Pharmaceuticals, Inc.; Eli Lilly and Company; EuroImmun; F. Hoffmann-La Roche Ltd and its affiliated company Genentech, Inc.; Fujirebio; GE Healthcare; IXICO Ltd.; Janssen Alzheimer Immunotherapy Research & Development, LLC.; Johnson & Johnson Pharmaceutical Research & Development LLC.; Lumosity; Lundbeck; Merck & Co., Inc.; Meso Scale Diagnostics, LLC.; NeuroRx Research; Neurotrack Technologies; Novartis Pharmaceuticals Corporation; Pfizer Inc.; Piramal Imaging; Servier; Takeda Pharmaceutical Company; and Transition Therapeutics. The Canadian Institutes of Health Research is providing funds to support ADNI clinical sites in Canada. Private sector contributions are facilitated by the Foundation for the National Institutes of Health (www.fnih.org). The grantee organization is the Northern California Institute for Research and Education, and the study is coordinated by the Alzheimer’s Therapeutic Research Institute at the University of Southern California. ADNI data are disseminated by the Laboratory for Neuro Imaging at the University of Southern California.

Additional information to include in an acknowledgment statement can be found on the LONI site: https://adni.loni.usc.edu/wp-content/uploads/how_to_apply/ADNI_Data_Use_Agreement.pdf.

##### FASe_Families (sa000004) data:

This work was supported by grants from the National Institutes of Health (R01AG044546, P01AG003991, RF1AG053303, R01AG058501, U01AG058922, RF1AG058501 and R01AG057777). The recruitment and clinical characterization of research participants at Washington University were supported by NIH P50 AG05681, P01 AG03991, and P01 AG026276. This work was supported by access to equipment made possible by the Hope Center for Neurological Disorders, and the Departments of Neurology and Psychiatry at Washington University School of Medicine.

We thank the contributors who collected samples used in this study, as well as patients and their families, whose help and participation made this work possible. This work was supported by access to equipment made possible by the Hope Center for Neurological Disorders, and the Departments of Neurology and Psychiatry at Washington University School of Medicine

##### KnightADRC (sa000008) data:

This work was supported by grants from the National Institutes of Health (R01AG044546, P01AG003991, RF1AG053303, R01AG058501, U01AG058922, RF1AG058501 and R01AG057777). The recruitment and clinical characterization of research participants at Washington University were supported by NIH P50 AG05681, P01 AG03991, and P01 AG026276. This work was supported by access to equipment made possible by the Hope Center for Neurological Disorders, and the Departments of Neurology and Psychiatry at Washington University School of Medicine.

We thank the contributors who collected samples used in this study, as well as patients and their families, whose help and participation made this work possible. This work was supported by access to equipment made possible by the Hope Center for Neurological Disorders, and the Departments of Neurology and Psychiatry at Washington University School of Medicine.

##### AMP-AD (sa000011) data:

Mayo RNAseq Study- Study data were provided by the following sources: The Mayo Clinic Alzheimer's Disease Genetic Studies, led by Dr. Nilufer Ertekin-Taner and Dr. Steven G. Younkin, Mayo Clinic, Jacksonville, FL using samples from the Mayo Clinic Study of Aging, the Mayo Clinic Alzheimer's Disease Research Center, and the Mayo Clinic Brain Bank. Data collection was supported through funding by NIA grants P50 AG016574, R01 AG032990, U01 AG046139, R01 AG018023, U01 AG006576, U01 AG006786, R01 AG025711, R01 AG017216, R01 AG003949, NINDS grant R01 NS080820, CurePSP Foundation, and support from Mayo Foundation. Study data includes samples collected through the Sun Health Research Institute Brain and Body Donation Program of Sun City, Arizona. The Brain and Body Donation Program is supported by the National Institute of Neurological Disorders and Stroke (U24 NS072026 National Brain and Tissue Resource for Parkinson's Disease and Related Disorders), the National Institute on Aging (P30 AG19610 Arizona Alzheimer's Disease Core Center), the Arizona Department of Health Services (contract 211002, Arizona Alzheimer's Research Center), the Arizona Biomedical Research Commission (contracts 4001, 0011, 05-901 and 1001 to the Arizona Parkinson's Disease Consortium) and the Michael J. Fox Foundation for Parkinson's Research

ROSMAP- We are grateful to the participants in the Religious Order Study, the Memory and Aging Project. This work is supported by the US National Institutes of Health [U01 AG046152, R01 AG043617, R01 AG042210, R01 AG036042, R01 AG036836, R01 AG032990, R01 AG18023, RC2 AG036547, P50 AG016574, U01 ES017155, KL2 RR024151, K25 AG041906-01, R01 AG30146, P30 AG10161, R01 AG17917, R01 AG15819, K08 AG034290, P30 AG10161 and R01 AG11101.

Mount Sinai Brain Bank (MSBB)- This work was supported by the grants R01AG046170, RF1AG054014, RF1AG057440 and R01AG057907 from the NIH/National Institute on Aging (NIA). R01AG046170 is a component of the AMP-AD Target Discovery and Preclinical Validation Project. Brain tissue collection and characterization was supported by NIH HHSN271201300031C.

##### UPitt Kamboh (sa000012) data:

This study was supported by the National Institute on Aging (NIA) grants AG030653, AG041718, AG064877 and P30-AG066468.

##### NACC Genentech (sa000013) data:

We would like to thank study participants, their families, and the sample collectors for their invaluable contributions. This research was supported in part by the National Institute on Aging grant U01AG049508 (PI Alison M. Goate). This research was supported in part by Genentech, Inc. (PI Alison M. Goate, Robert R. Graham).

The NACC database is funded by NIA/NIH Grant U01 AG016976. NACC data are contributed by these NIA-funded ADCs: P30 AG013846 (PI Neil Kowall, MD), P50 AG008702 (PI Scott Small, MD), P50 AG025688 (PI Allan Levey, MD, PhD), P30 AG010133 (PI Andrew Saykin, PsyD), P50 AG005146 (PI Marilyn Albert, PhD), P50 AG005134 (PI Bradley Hyman, MD, PhD), P50 AG016574 (PI Ronald Petersen, MD, PhD), P30 AG013854 (PI M. Marsel Mesulam, MD), P30 AG008017 (PI Jeffrey Kaye, MD), P30 AG010161 (PI David Bennett, MD), P30 AG010129 (PI Charles DeCarli, MD), P50 AG016573 (PI Frank LaFerla, PhD), P50 AG005131 (PI Douglas Galasko, MD), P30 AG028383 (PI Linda Van Eldik, PhD), P30 AG010124 (PI John Trojanowski, MD, PhD), P50 AG005142 (PI Helena Chui, MD), P30 AG012300 (PI Roger Rosenberg, MD), P50 AG005136 (PI Thomas Grabowski, MD), P50 AG005681 (PI John Morris, MD), P30 AG028377 (Kathleen Welsh-Bohmer, PhD), and P50 AG008671 (PI Henry Paulson, MD, PhD).

Samples from the National Cell Repository for Alzheimer’s Disease (NCRAD), which receives government support under a cooperative agreement grant (U24 AG21886) awarded by the National Institute on Aging (NIA), were used in this study. We thank contributors who collected samples used in this study, as well as patients and their families, whose help and participation made this work possible.

The Alzheimer's Disease Genetics Consortium supported the collection of samples used in this study through National Institute on Aging (NIA) grants U01AG032984 and RC2AG036528.

##### CacheCounty (sa000014) data:

We acknowledge the generous contributions of the Cache County Memory Study participants. Sequencing for this study was funded by RF1AG054052 (PI: John S.K. Kauwe).

##### PSP-NIH-CurePSP-Tau (sa000015) data:

This project was funded by the NIH grant UG3NS104095 and supported by grants U54NS100693 and U54AG052427. Queen Square Brain Bank is supported by the Reta Lila Weston Institute for Neurological Studies and the Medical Research Council UK. The Mayo Clinic Florida had support from a Morris K. Udall Parkinson's Disease Research Center of Excellence (NINDS P50 #NS072187), CurePSP and the Tau Consortium. The samples from the University of Pennsylvania are supported by NIA grant P01AG017586.

##### PSP-CurePSP-Tau (sa000016) data:

This project was funded by the Tau Consortium, Rainwater Charitable Foundation, and CurePSP. It was also supported by NINDS grant U54NS100693 and NIA grants U54NS100693 and U54AG052427. Queen Square Brain Bank is supported by the Reta Lila Weston Institute for Neurological Studies and the Medical Research Council UK. The Mayo Clinic Florida had support from a Morris K. Udall Parkinson's Disease Research Center of Excellence (NINDS P50 #NS072187), CurePSP and the Tau Consortium. The samples from the University of Pennsylvania are supported by NIA grant P01AG017586. Tissues were received from the Victorian Brain Bank, supported by The Florey Institute of Neuroscience and Mental Health, The Alfred and the Victorian Forensic Institute of Medicine and funded in part by Parkinson’s Victoria and MND Victoria. We are grateful to the Sun Health Research Institute Brain and Body Donation Program of Sun City, Arizona for the provision of human biological materials (or specific description, e.g. brain tissue, cerebrospinal fluid). The Brain and Body Donation Program is supported by the National Institute of Neurological Disorders and Stroke (U24 NS072026 National Brain and Tissue Resource for Parkinson's Disease and Related Disorders), the National Institute on Aging (P30 AG19610 Arizona Alzheimer's Disease Core Center), the Arizona Department of Health Services ( contract 211002, Arizona Alzheimer's Research Center), the Arizona Biomedical Research Commission (contracts 4001, 0011, 05-901 and 1001 to the Arizona Parkinson's Disease Consortium) and the Michael J. Fox Foundation for Parkinson's Research. Biomaterial was provided by the Study Group DESCRIBE of theClinical Research of the German Center for Neurodegenerative Diseases (DZNE).

##### PSP_UCLA (sa000017) data:

If data are used for a publication, “on behalf of the AL-108-231 investigators” should be included in the authorship list.

#### ADSP R3 Cohorts

##### sa000001- Alzheimer's Disease Sequencing Project (ADSP)

###### ADSP Discovery WGS

The initial phase of the ADSP research plan is called the Discovery Phase. Samples were selected from well-characterized study cohorts of individuals with or without an AD diagnosis and the presence or absence of known risk factor genes. The ADSP generated three sets of genome sequence data for these samples as part of the Discovery Phase: (1) WGS for 584 samples from 113 multiplex families (two or more affected per family), (2) Whole Exome Sequence (WES) for 5,096 AD cases and 4,965 controls, and (3) WES of an Enriched sample set comprised of 853 AD cases from multiply affected families and 171 Hispanic controls. The Case-Control and Enriched Case Study spans 24 cohorts provided by the Alzheimer’s Disease Genetics Consortium (ADGC) and the Cohorts for Heart and Aging Research in Genomic Epidemiology (CHARGE) Consortium.

###### ADSP Discovery Family-Based Extension

To further assess the genomes in multiply affected families, under funding provided by NHGRI, an additional 427 samples were whole genome sequenced. This included 107 additional samples from families studied under the Discovery Phase, 175 samples from 47 new families, and 145 Hispanic Controls. This portion of the study is called the Discovery Extension Phase. The Family Based Study spans seven cohorts provided by the Alzheimer’s Disease Genetics Consortium (ADGC) and the Cohorts for Heart and Aging Research in Genomic Epidemiology (CHARGE) Consortium.

###### The ADSP Discovery Case-Control Based Extension

Under funding provided by NHGRI, an additional 3,000 subjects were whole genome sequenced. This included 1,466 cases and 1,534 controls. Of these 1,000 each of Non-Hispanic White (NHW), Caribbean Hispanic (CH), and African American (AA) descent were sequenced. Of these a total of 739 autopsy samples were sequenced [568 cases (500 NHW cases and 68 AA cases) and 171 controls (164 NHW and 7 AA)]. The Case-Control and Enriched Case Study spans 24 cohorts provided by the Alzheimer’s Disease Genetics Consortium (ADGC) and the Cohorts for Heart and Aging Research in Genomic Epidemiology (CHARGE) Consortium.

###### ADSP-FUS1

The ADSP-FUS is a National Institute on Aging (NIA) initiative focused on identifying genetic risk and protective variants for late-onset Alzheimer Disease (LOAD). A concern in AD genetic studies is a lack of racial-ethnic diversity. The ADSP-FUS collects and sequences existing ethnically diverse and unique cohorts with clinical data to expand the utility of new discoveries for individuals from all populations. The first release of FUS data 3,250 AD cases, 4,149 cognitively normal individuals, 194 individuals with mild cognitive impairment, and 567 with unknown or other dementia from seven datasets (PR1066, ADC Autopsy, ADGC African American (release 1), HIHG Brain Bank, ADNI-WGS-2, APOE Extremes, and StEP AD). Counts below may not be the final number of genomes released in this dataset, but instead what were sent for sequencing. Brief descriptions of the individual datasets are provided below. Detailed descriptions of the datasets, including methods used for their adjudication can be found in the datasets README files located in ADSP-FUS1-WGS-StudyDescriptions.zip.

**The Puerto Rican 10/66 Study (PR1066)** (https://www.alz.co.uk/1066/), is an Alzheimer’s Disease International study of dementia in Puerto Rico that began in 2007 (Dr. Ivonne Jimenez-Velazquez, PI). Individuals were recruited as part of the the 10/66 Population Based Study of Dementia using standard protocols. As part of this study, sociodemographic information and detailed clinical history of memory decline were collected. In addition, the Clinical Dementia Rating scale (CDR), and Community Screening Interview for Dementia, and Petersen ADL criteria were collected for all individuals. Neurocognitive testing (CERAD battery) was available for some participants. Participants were adjudicated for dementia. Lastly, the total number of samples that were whole genome sequenced is 1,565 with the breakdown of 140 Cases; 1,245 Controls and 180 MCI’s.

Prince M, Ferri CP, Acosta D, et al. The protocols for the 10/66 dementia research group population- based research programme. BMC Public Health. 2007;7(1):165.

**The Alzheimer’s Disease Center Autopsy (ADC) Cohort** includes 1,500 cases and 1,372 controls from the National Institute on Aging’s ADC’s (https://www.nia.nih.gov/health/alzheimers-disease-research-centers). The total number of samples that were whole genome sequenced was 2,845. This cohort has autopsy reports available on (651 cases & 32 controls) and only clinical data available on (849 cases & 1,320 controls).

These individuals are well phenotyped and will help to increase the currently underpowered NHW sample with WGS data from the ADSP Discovery and Discovery Extension Phases (1,212 cases and 524 controls).

**The ADGC African American Dataset (release 1).** The ADGC AA release 1 dataset consists of 1,923 individuals collected as part of the Alzheimer Disease Genetics Consortium. This release consists of two datasets, a dataset of 342 cases, 655 controls, and 29 unknown/other dementia from the ADC’s, and a dataset of 350 cases, 544 controls, and 3 unknown/other dementia collected by investigators at the University of Miami, Case Western Reserve University, and North Carolina A&T.

**John P. Hussman Institute for Human Genomics (HIHG) Brain Bank.** The HIHG Brain, Bank autopsy individuals followed the typical clinical course of disease. This sample is a clinic and community outreach sample ascertained in North and South Carolina and Virginia. The total number of samples that were whole genome sequenced is 96 with available autopsy reports for the breakdown of 92 Cases and 4 Controls (cases include AD and other related dementias).

**ADNI-WGS-2.** The Alzheimer’s Disease Neuroimaging Initiative (ADNI) is a longitudinal multicenter study designed to develop clinical, imaging, genetic, and biochemical biomarkers for the early detection and tracking of Alzheimer’s disease (AD). Since its launch in 2004, the landmark public-private partnership has made major contributions to AD research, enabling the sharing of data between researchers around the world.

In 2019, additional 769 ADNI participants underwent whole genome sequencing as a collaboration between ADNI and ADSP Follow-Up Study (FUS). WGS was performed at the the American Genome Center Core Laboratory at the Uniformed Services University of the Health Sciences, Bethesda, MD, directed by Clifton L Dalgard, PhD, CLS. This included 238 with AD dementia, 320 with MCI, 105 with SMC (significant memory concerns without cognitive impairment), and 106 cognitively unimpaired controls at baseline. Following initial quality control, data from 12 individuals was excluded. For more information about the ADNI study and additional samples sequenced, see the ADNI study page snd000002.

**APOE Extremes.** The APOE extremes study entails Alzheimer’s disease (AD) case-control association analysis using an age extremes sampling approach stratified by APOE genotype, comparing younger onset AD cases against older cognitively normal controls. We queried the National Alzheimer’s Coordinating Center database to select individuals of any ancestry that lacked sequencing data and matched the following criteria: APOE ε4/ε4 or ε3/ε4 AD cases with age at onset ≤ 65 years, APOE ε3/ε3 AD cases with age at onset ≤ 70 years, APOE ε4/ε4 controls with age at last assessment ≥ 75 years, APOE ε3/ε4 controls with age at last assessment ≥ 80 years, or APOE ε3/ε3 controls with age at last assessment ≥ 85 years. DNA samples were provided by the National Cell Repository of Alzheimer’s Disease. Whole genome sequencing was performed at the American Genome Center, Uniformed Services University of the Health Sciences. PCR-free, paired-end Illumina TruSeq libraries were generated then underwent paired-end sequencing on an Illumina HiSeq X Ten (v2.5 chemistry) using 150 bp, paired-end cycles.

Chia R, et al. Genome sequencing analysis identifies new loci associated with Lewy body dementia and provides insights into its genetic architecture. Nat Genet. 2021 Feb 15. doi: 10.1038/s41588-021-00785-3. Online ahead of print. https://pubmed.ncbi.nlm.nih.gov/33589841.

**Grant Funding**

1.	National Institute on Aging U01AG049508 “Modifier Genes that Influence Age at Onset or Protect Against Development of Alzheimer's Disease” (PI Alison M. Goate).

2.	National Institute on Aging U01AG052411 “Identification and Characterization of AD Risk Networks Using Multi-dimensional “Omics” Data” (PI Alison M. Goate, Carlos Cruchaga, Bin Zhang).

3.	National Institute of Neurological Disorders and Stroke Intramural Research Program 1ZIANS003154 (PI Sonja W. Scholz).

4.	National Institute on Aging Intramural Research Program 1ZIAAG000935 (PI Bryan J. Traynor).

5.	Department of Defense HU0001-18-2-0038 (PI Clifton L. Dalgard).

6.	National Heart, Lung, and Blood Institute IAA-A-HL-007.001 (PI Clifton L. Dalgard).

**Principal Investigators**- Alison M. Goate, Carlos Cruchaga, Bin Zhang, Sonja W. Scholz, Bryan J. Traynor, J. Raphael Gibbs, Clifton L. Dalgard.

**Publication Acknowledgments**

We would like to thank study participants, their families, and the sample collectors for their invaluable contributions. We would like to thank J. Raphael Gibbs for undertaking whole genome sequence data processing. This research was supported in part by National Institute on Aging grants U01AG049508 (PI Alison M. Goate) and U01AG052411 (PI Alison M. Goate, Carlos Cruchaga, Bin Zhang). This research was supported in part by the Intramural Research Program of the National Institutes of Health (National Institute on Aging, National Institute of Neurological Disorders and Stroke; project numbers: 1ZIAAG000935 [PI Bryan J. Traynor], 1ZIANS003154 [PI Sonja W. Scholz]). The American Genome Center is supported by the Department of Defense award: HU0001-18-2-0038 (PI Clifton L. Dalgard) and in part by a National Heart, Lung, and Blood Institute grant: IAA-A-HL-007.001 (PI Clifton L. Dalgard). The opinions and assertions expressed herein are those of the author(s) and do not necessarily reflect the official policy or position of the Uniformed Services University or the Department of Defense, or the Henry M. Jackson Foundation for the Advancement of Military Medicine, Inc. or the U.S. Government. The American Genome Center receives administrative and programmatic support from the Henry Jackson M. Foundation for the Advancement of Military Medicine.

Stanford Extreme Phenotypes in AD (StEP AD). The overall goals of the Stanford Extreme Phenotypes in AD (StEP AD) project (Dr. Michael Greicius) are to identify and characterize novel genetic variants that promote resilience to AD pathology in the presence of the APOE4 allele or that drive pathogenesis in the absence of the APOE4 allele. Genomes are collected from several sources, some intramural and some extramural. Invariably, the cognitive assessment protocols for these different sources vary somewhat but all include APOE genotyping, extensive neuropsychological testing, collection of one or more AD biomarkers, and consensus adjudication.

Genomes were sequenced for subjects in the following three categories:

1. Protected APOE4 carriers that have the APOE3/4 genotype, are at least 80 years old, and have normal cognition. If additional follow-up is expected we will accept subjects as young as 77.

2. Super-protected APOE4 carriers that have the APOE4/4 genotype, are at least 70 years old, and have normal cognition. If additional follow-up is expected we will accept subjects as young as 67.

3. APOE4-negative, early-onset cases that have the APOE2/2, 2/3, or 3/3 genotype and are diagnosed with probable AD before age 65. Most are also negative for known PSEN1, PSEN2 or APP mutations.

Funding for this project comes from the NIA (1R01AG060747).

##### sa000002- Alzheimer's Disease Neuroimaging Initiative (ADNI)

In 2012, the Brin-Wojcicki Foundation and the Alzheimer’s Association donated funds to support whole genome sequencing (WGS) of 818 ADNI participants (at baseline: 48 with AD, 484 with MCI, and 286 cognitively unimpaired controls). Samples were sent to Illumina, where non-CLIA WGS, as well as Illumina Omni 2.5M genome-wide association study (GWAS) single nucleotide polymorphism (SNP) arrays, were performed on each sample and completed in 2013. Basic quality control checks were then performed and additional checks were performed, dropping 9 individuals. Further details of Genetics Core studies and datasets from the ADNI-1/GO/2 phases can be found here: https://www.ncbi.nlm.nih.gov/pmc/articles/PMC4510473/.

In 2019, additional 769 ADNI participants underwent whole genome sequencing as a collaboration between ADNI and ADSP Follow-Up Study (FUS). WGS was performed at the the American Genome Center Core Laboratory at the Uniformed Services University of the Health Sciences, Bethesda, MD, directed by Clifton L Dalgard, PhD, CLS. This included 238 with AD dementia, 320 with MCI, 105 with SMC (significant memory concerns without cognitive impairment), and 106 cognitively unimpaired controls at baseline. Following initial quality control, data from 12 individuals was excluded. These samples are part of the ADSP-FUS1-WGS sample set mentioned above (ADNI-WGS-2).

##### sa000004- The Familial Alzheimer Sequencing Project (FASe)

GWAS studies were very successful in identifying genetic loci associated with AD risk. However, these studies could not point to the actual causal variant. In this study, WES and/or WGS data is being generated from families densely affected by Alzheimer disease (AD) under the hypothesis that these families will be enriched for rare genetic variants that confer risk to AD. This study included late onset families with a high proportion of AD cases (at least three affected members with DNA data available). The families originated from the NCRAD, NIA-LOAD, Knight-ADRC and DIAN-EXR cohorts. The proband of the family could not be a carrier of known pathogenic mutations in the Mendelian genes for AD (APP, PSEN1, PSEN2) or Frontotemporal Dementia (GRN, MAPT, C9ORF72). Cases within families had to be 65 years old or older at age at onset (AAO) and have a CDR >=0.5; controls within families had to have a CDR=0 at last assessment and be older than the largest AAO within the family. AD definition was based on a combination of both clinical and pathological information if available. Clinical diagnosis was overruled by pathological diagnosis. Subjects are Non-Hispanic white from North America, based on self-reporting. Autopsy Information was provided if available, but it was not a requirement for enrollment.

##### sa000008- Charles F. and Joanne Knight Alzheimer’s Disease Research Center (KnightADRC)

The search for novel risk factors for Alzheimer disease relies on access to accurate and deeply phenotyped datasets. The Memory and Aging Project at the Knight-ADRC (Knight ADRC-MAP) collects plasma, CSF, fibroblast, neuroimaging, clinical and cognition data longitudinally and autopsied brain samples. We are using multi-tissue (brain, CSF and plasma) multi-omic data (genetics, epigenomics, transcriptomics, proteomics and metabolomics) to identify novel risk and protective variants, create new prediction models and identify drug targets. The study cohort includes MAP participants from the Knight-ADRC at Washington University in St. Louis (MO). MAP participants have to be at least 65 years old and have no memory problems or mild dementia at the time of enrollment. There is no age at onset criteria for this cohort. Cases had to have a CDR >=0.5 whereas controls had to have a CDR=0 at last assessment. AD definition is based on a combination of both clinical and pathological information if available. Pathologic diagnosis will overrule clinical diagnosis. Participants are Non-Hispanic white from North America (95%) and African American (5%). Autopsy information was provided if available, but it is not a requirement for enrollment.

##### sa000011- Accelerating Medicines Partnership-Alzheimer’s Disease (AMP-AD)

The Accelerating Medicines Partnership- Alzheimer’s Disease Target Discovery and Preclinical Validation (AMP-AD) has supported the generation of whole genome data from three studies: the MAYO RNAseq Study, the Mount Sinai Brain Bank, and the ROSMAP study. The Genome Center for AD (GCAD) has partnered with the investigators of these studies to generate a joint genotype called dataset consisting of the AMP-AD genomes along with the ADSP and other Alzheimer's Disease and Related Dementia projects. Data were obtained directly from each AMP-AD investigator and harmonized by the Genome Center for AD on the VCPA1.1 pipeline. Additional information about each of the three studies can be obtained from the AMP-AD Knowledge Portal.

##### sa000012- University of Pittsburg- Kamboh (UPitt)

The project goal is to perform whole-genome sequencing (WGS) on Alzheimer’s disease (AD) cases to identify both common and uncommon variants associated with AD-related phenotypes (e.g., disease progression, survival time). All subjects were Caucasians with probable or possible having age at onset of 60 or above. Whole-genome sequencing was performed using Illumina HiSeqX.

##### sa000013- NACC Genentech Study

The APOE extremes study entails Alzheimer’s disease (AD) case-control association analysis using an age extremes sampling approach stratified by APOE genotype, comparing younger onset AD cases against older cognitively normal controls. We queried the National Alzheimer’s Coordinating Center database to select individuals of self-reported European ancestry that lacked sequencing data and matched the following criteria: APOE ε4/ε4 or ε3/ε4 AD cases with age at onset ≤ 65 years, APOE ε4/ε4 controls with age at last assessment ≥ 75 years, or APOE ε3/ε4 controls with age at last assessment ≥ 80 years. DNA samples were provided by the National Cell Repository of Alzheimer’s Disease. Whole genome sequencing was performed at Illumina. PCR-amplified, paired-end Illumina libraries were generated then underwent paired-end sequencing on an Illumina HiSeq 2000.

##### sa000014- Cache County Study

Investigators for this study report the development and use of an innovative, powerful approach to identify functional variants that provide AD resilience to high-risk individuals. In this study, pedigrees were identified with a statistical excess of AD mortality that also included at least four AD high-risk resilient individuals. Linkage analysis was performed in these families and whole genome sequence (WGS) data from resilient individuals was used to interrogate identified linkage regions for candidate variants. Finally, RAB10 and SAR1A was tested for biological impact in vitro. The results of this study suggest that RAB10 variants impact risk for AD and that RAB10 may represent a promising therapeutic target for AD prevention.

Researchers use the Utah Population Database (UPDB) to identify large pedigrees with an evidence of excess AD mortality (i.e., families with a higher number of AD deaths than expected). Pedigrees with an excess of AD deaths were identified by comparing the observed (i.e., number of affected individuals in the pedigree) to the expected numbers of AD-affected individuals within the pedigree. The expected number of AD deaths was estimated using population-based, cohorts specific rates of AD death estimated from all Utah death certificates for individuals in the UPDB genealogy.

“AD resilient individuals” defined as individuals who are at least 75 years old, cognitively normal, and carry at least one APOE ε4 allele were included in this study. AD deaths were identified by the inclusion of ICD codes for AD death in individual death certificates, where deaths were only considered an AD death if the death certificate included AD ICD codes (ICD9 331.0; ICD10 F00 or G30) as a primary or contributing cause-of-death.

An expert panel of neurologists, neuropsychologists, neuropsychiatrists, and a cognitive neuroscientist assigned final diagnoses of dementia following standard research protocols (e.g., NINCDS-ADRDA criteria for AD or NINCDS-AIREN criteria for vascular dementia).

##### sa000015- NIH, CurePSP and Tau Consortium PSP WGS

Progressive supranuclear palsy (PSP) is the most common frontotemporal lobar degeneration associated with tau pathology. The majority of patients with PSP can be accurately diagnosed antemortem due to a characteristic clinical syndrome; however, there are atypical clinicopathologic forms of PSP that can mimic Parkinson’s disease, frontotemporal dementia, progressive nonfluent aphasia and corticobasal syndrome. While rare pathogenic variants, common risk factors, and – more recently – rare risk-associated variants have been identified in PSP, a significant proportion of the heritability for neurodegenerative tauopathies and other frontotemporal lobar degenerations remains unexplained, strongly suggesting that additional genetic risk factors await discovery. We assembled PSP samples for whole-genome sequencing (WGS) of neuropathologically characterized PSP, with the majority coming from the Mayo Clinic in Jacksonville. Other major contributors were the University of Pennsylvania and University College London.

The SeqLab Core at the Henry M. Jackson Foundation for the Advancement of Military Medicine, Inc., Uniformed Services University of the Health Sciences (HJF/USU) conducted genomic sequencing of de-identified DNA samples. Library preparation used the Illumina PCR-free DNA workflow on SeqLab automated Hamilton STAR liquid handling platforms. Clustered libraries were sequenced using the Illumina HiSeq X platform. FASTQ data were generated and genomic variants called after quality control, alignment and calling performed on HAS 2.0 software algorithm. The Genome Center for Alzheimer’s Disease (GCAD), at the University of Pennsylvania, processed the sequence data using a standardized pipeline.

##### sa000016- CurePSP and Tau Consortium PSP WGS

Progressive supranuclear palsy (PSP) is the most common frontotemporal lobar degeneration associated with tau pathology. The majority of patients with PSP can be accurately diagnosed antemortem due to a characteristic clinical syndrome; however, there are atypical clinicopathologic forms of PSP that can mimic Parkinson’s disease, frontotemporal dementia, progressive nonfluent aphasia and corticobasal syndrome. While rare pathogenic variants, common risk factors, and – more recently – rare risk-associated variants have been identified in PSP, a significant proportion of the heritability for neurodegenerative tauopathies and other frontotemporal lobar degenerations remains unexplained, strongly suggesting that additional genetic risk factors await discovery. We assembled 900 PSP samples for whole-genome sequencing (WGS) of neuropathologically characterized PSP. About half of the samples come from the Mayo Clinic in Jacksonville, and the others were compiled from investigators across the US and Europe. Macrogen performed HiSeq whole genome sequencing. The Genome Center for Alzheimers Disease (GCAD) process the files with their standardized pipeline.

##### sa000017- UCLA Progressive Supranuclear Palsy

This study includes subjects with Progressive Supranuclear Palsy (PSP). Subjects were enrolled in the davenutide PSP Phase Trail 2/3. Additional subjects were clinically diagnosed PSP at the University of California, San Francisco Memory and Aging Center.

Participants met the modified Neuroprotection and Natural History in Parkinson Plus Syndrome study criteria for PSP.

See https://clinicaltrials.gov/ct2/show/NCT01110720 for comprehensive inclusion and exclusion criteria.
